# Single-cell splicing analysis with *ISSAC* links cell type-specific and cell state-dependent sQTLs to neurological disorders

**DOI:** 10.64898/2026.05.06.26352548

**Authors:** Yuntian Zhang, Wenjing Wang, Zhi Yang Tan, Yihan Tong, Chang Xu, Anjing Liu, Donald Sim, Shuyi Jin, Chi Tian, Natacha Comandante Lou, Xavier Roca, Philip L. De Jager, David A. Benett, The Alzheimer’s Disease Functional Genomics Consortium, Gao Wang, Boxiang Liu

## Abstract

Single-cell RNA sequencing enables comprehensive profiling of gene expression and splicing at cellular resolution, revealing cell type-specific and cell state-dependent regulation (variation within cell types based on their functional states). While genetic studies of expression (eQTLs) in single cells are well established, the genetic regulation of alternative splicing in single cells remains challenging. Existing single-cell splicing QTL (sQTL) studies perform pseudobulk aggregation using bulk analysis methods, which reduces power to detect cell type-specific sQTLs and cannot capture cell state-dependent splicing regulation. Here, we introduce *ISSAC* to directly quantify metacell-level splice site usage and map cell type- and cell state-specific sQTLs through generalized linear mixed models. In real-world benchmarking on peripheral blood single-cell data, *ISSAC* identified 1.4- to 2.5-fold more cell type-specific sQTLs than pseudobulk sQTL analysis, and uniquely enabled cell state-dependent sQTL discovery. We applied *ISSAC* to a harmonized aging brain resource consisting of approximately 3 million dorsolateral prefrontal cortex (DLPFC) single nuclei from 722 donors. *ISSAC* identified 31,318 independent *cis*-sQTLs across seven major cell types and 16,861 independent *cis*-sQTLs across 67 subcell types, with ∼67% of sGenes showing no overlap with eGenes. We identified 369 independent sQTLs whose genetic effects were mediated by various cell states such as dendrite development and synaptic signaling. Additionally, we uncovered 194 Alzheimer’s-biased and 207 sex-biased sGenes, as well as 142 risk genes that colocalized with neurological disorders including Alzheimer’s disease, Neuroticism, Amyotrophic lateral sclerosis, Parkinson’s disease, Lewy body dementia and Schizophrenia. Specifically, we functionally validated a causal variant rs11549690 modulating *TRPT1* exon 7 skipping to influence neuroticism risk.

## Introduction

Alternative RNA splicing is a crucial mediator of complex diseases^1-5^. Splicing quantitative trait loci (sQTLs) are genetic variants regulating alternative splicing, thereby affecting the relative abundance of transcript isoforms^6^. Studies have identified tens of thousands of tissue-specific sQTLs across a wide range of human tissues^7,8^, yet tissue-level analyses inherently miss cell type-specific genetic regulation. To address this gap, recent studies have mapped cell type-specific effects with bulk RNA-seq on cell cultures^4,9,10^, FACS-sorted primary cells^11-14^, as well as with single-cell RNA-seq data (scRNA-seq)^15^. In particular, the increasing scale and comprehensive cell type coverage offered by single-cell RNA-seq has made it a popular choice for recent cell type-specific sQTL studies^15-18^.

However, current single-cell sQTL studies must rely on pseudobulk analysis^15^ because existing sQTL mapping tools are designed for bulk RNA-seq data^5,10,19-21^. These approaches face critical limitations from both molecular phenotype profiling and statistical analysis prospectives. First, pseudobulk aggregation misses important information about cell-to-cell variability. It is widely accepted that, within the same cell type, cell state is a key determinant of genetic regulation^22,23^. However, neither cell culture, FACS-sorted, nor pseudobulk analysis provides information about cell state-dependent regulation. Second, single-cell eQTL studies have shown that each single cell can be treated as a data point, and proper statistical modeling of repeated single-cell measurements can boost statistical power for eQTL calling^24^. However, the current pseudobulk sQTL procedure does not properly account for the repeated single-cell measurements within the same donor, which leads to a reduction in power. Third, current isoform-^5,10,20^, exon-^21^, and intron-based^19^ bulk-level sQTL mapping assumes even RNA-seq read coverage typical for bulk RNA-seq. In contrast, scRNA-seq data often violates this assumption with 5’/3’-biased read coverage^25-27^, and it is unclear whether isoform-, exon-, and intron-based splicing phenotypes are appropriate for end-biased read coverage. In our recent scRNA-seq sQTL study, pseudobulk analysis missed intron retention events as a key component of splicing regulation^15^.

To address these limitations, we introduce *ISSAC* (<u>I</u>ntegrative <u>s</u>ingle-cell <u>s</u>plicing <u>a</u>nalysis and QTL <u>c</u>aller), a scalable method that quantifies splice site usage and maps sQTL using a generalized linear mixed model in single-cell or single-nucleus RNA-seq data. To address phenotype profiling challenges, *ISSAC* directly quantifies splice site usage, a metric not only robust to 5’/3’ read bias but also enables detection of both canonical splicing and intron retention events missed by intron-based methods^19^. Previous work has also explored splice site usage as a quantitative phenotype in bulk RNA-seq cohorts^28,29^. Furthermore, *ISSAC* integrates neighboring cells with similar gene expression profiles into metacells, maintaining cell-to-cell variability critical for detecting cell state-dependent regulation while mitigating sparsity in single-cell data^30^. To properly model this metacell structure and maximize statistical power, *ISSAC* employs a generalized mixed-effect model that treats splice site usage as a binomial random variable, and accounts for repeated metacell measurements from the same individual^31^.

Through comprehensive simulated and real-world data benchmarks, we demonstrated that *ISSAC* outperforms traditional bulk-based methods in statistical power, while uniquely enabling cell state-dependent sQTL discovery. We applied *ISSAC* to map cell type-specific and cell state-dependent sQTLs in the human dorsolateral prefrontal cortex (DLPFC), an area in the human brain with particularly high isoform diversity^32^. Although studies have identified cell type-specific eQTLs^33^, there has been minimal investigation on the cell type-specific sQTLs in DLPFC regions. In this study, we aggregated 3 million single-nucleus profiles from two DLPFC resources consisting of 424 and 298 donors, respectively^34,35^. We identified 31,318 independent *cis*-sQTLs across seven main cell types and an additional 16,861 independent *cis*-sQTLs across 67 subcell types, including those affecting intron retention that were missed by previous pseudobulk sQTL analyses. Among the seven major cell types, we discovered 399 AD-biased and 463 sex-biased sQTLs, highlighting abundant context-biased regulation of alternative splicing^36^. In addition, we identified 369 cell state-dependent sQTLs associated with dendrite development, synaptic signaling transmission, neuro-immunity, and other biological processes. Through colocalization and Mendelian randomization, we prioritized 142 sGenes that putatively mediates GWAS risk loci from six neurological disorders, including *MAPT* (excitatory neuron-specific, linked to both AD and PD), *GNL3* (Schizophrenia) and *TTC19* (Parkinson’s disease). Particularly, we functionally validated the genetic effect of a putative causal variant rs11549690 that modulated Neuroticism risk by affecting skipping of *TRPT1* exon 7.

## Results

### 1) Overview of *ISSAC*

*ISSAC* (<u>I</u>ntegrative <u>s</u>ingle-cell <u>s</u>plicing <u>a</u>nalysis and QTL <u>c</u>aller) is an integrated suite comprising three components: metacell generation, splice site usage quantification and sQTL mapping via a binomial mixed model. Unlike isoform-based methods^5,20^, *ISSAC* does not require transcript reference, thus enabling discovery of novel cell type-specific splicing events. Unlike intron-based methods such as LeafCutter^19^, *ISSAC* avoids intron clustering, which is often challenged by uneven read coverage^15^.

In brief, *ISSAC* first generates metacells by clustering cells from each donor based on PC embeddings of gene expression, using *k*-nearest neighbor graphs^37^ and Louvain clustering^38^ to group similar cells while preserving cell state heterogeneity (**Fig. 1a**). Next, to estimate splice site usage ratio, *ISSAC* quantifies junction reads by collapsing PCR-duplicates using cell barcode and UMI, retaining each unique junction-barcode-UMI combination only once to mitigate high PCR amplification biases in single-cell data (**Fig. 1b**). For each splice site, the usage ratio is defined as the proportion of junction reads spanning that site relative to junction reads from its “splicing partners”, *i.e*., nearby sites that join with the target site to form introns supported by junction reads (**Fig. 1c, d; Extended Data Fig. 1a**). This site-centric formulation of the splice site usage ratio (SSUR) offers several advantages over the intron usage ratio (IUR) employed by existing tools^19^. First, by restricting the denominator to junction reads from directly competing splicing partners at a given site, SSUR captures only biologically meaningful competition between alternative splice sites, whereas IUR aggregates all introns within a cluster – including distant introns that do not directly compete with the target site – potentially diluting the signal of local splicing changes. Second, because SSUR is anchored to individual splice sites rather than intron clusters, it naturally captures intron retention events: a reduction in junction-supporting reads relative to total overlapping reads at a given site is a direct signature of retention or incomplete splicing, whereas IUR cannot detect intron retention events, as retained introns produce no split reads. To benchmark the robustness of SSUR against IUR, we evaluated both metrics using simulated 10x Chromium 5’ and 3’ scRNA-seq data (**Supplementary Note 2.3**). SSUR were markedly less prone to 5’/3’ read bias inherent to 5’/3’ scRNA-seq library preparation than IUR, supporting SSUR’s adoption as the primary splicing quantification in *ISSAC* (**Supplementary Fig. 1**). For sQTL mapping, *ISSAC* fits a null binomial mixed-effect model incorporating fixed effects (sex, age, genotype PC, and hidden factors^39,40^ derived from molecular phenotypes) and random effects via a genetic relationship matrix (GRM) that accounts for repeated metacells per donor as well as potential genetic relatedness across donors, eliminating the needs to exclude related individuals from population-based cohorts (**Fig. 1e**). Following from previous mixed model approaches^31,41^, *ISSAC* uses penalized quasi-likelihood (PQL) with REML for parameter estimation, employing preconditioned conjugate gradient (PCG) approximation to avoid computationally intensive matrix inversion for under the REML framework^42^, as well as a sparse GRM to reduce memory usage. Score tests then evaluate per-variant association with splicing, with effect size defined as the regression slope between genotype and null-model adjusted phenotype. Further implementation details of each *ISSAC* module are provided in the **Methods** section.

**Fig 1:**
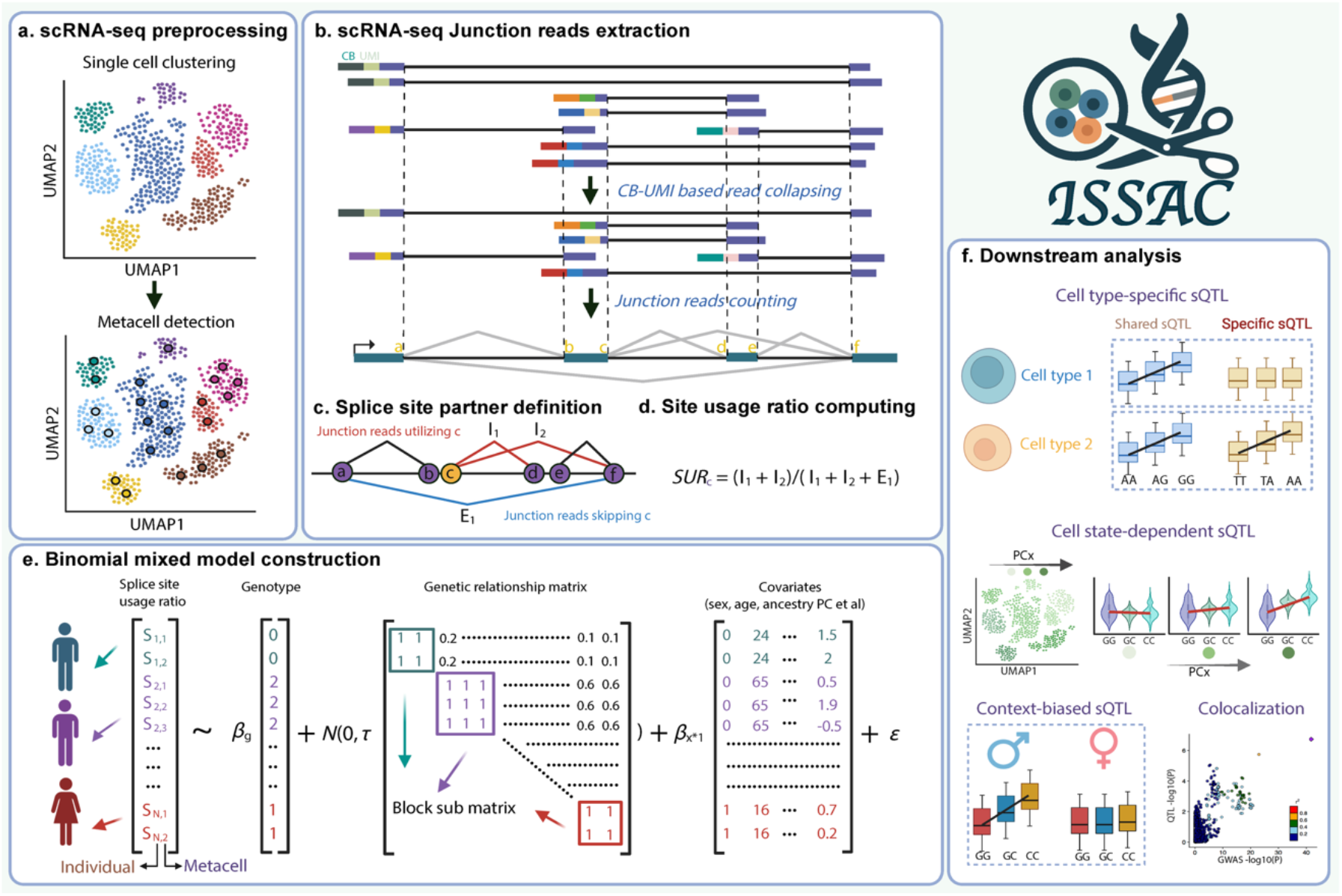
Schematics of *ISSAC* sQTL mapping procedures. **(a)** scRNA-seq preprocessing: After cell type annotation based on gene expression, *ISSAC* divides cells of one individual into several metacells per cell type based on principal components embeddings obtained from scRNA-seq gene expression matrix. **(b)** scRNA-seq Junction reads extraction: *ISSAC junctools* and *juncstats* modules enable extracting UMI-based junction read counts from aligned RNA-seq files. **(c)** Splice site partner definition: *ISSAC* identifies junction reads supporting the usage of the splice site c versus junction reads competing for its usage. **(d)** Splice site usage ratio computing: The numerator is defined as the sum of junction reads utilizing site c (I_1_ + I_2_) and the denominator is defined as the sum of supporting junction reads (I_1_ + I_2_) and junction reads competing the usage of site c (E_1_). **(e)** Binomial mixed model for association testing: *ISSAC* fits a null binomial mixed model by estimating parameters for fixed effects such as sex, age, ancestry PCs, and variance components for random effects represented by genetic relationship matrix; then performs single-variant score tests between genotype and splice site usage ratio to identify sQTLs. **(f)** Downstream applications: *ISSAC* enables cell type-specific sQTLs detection, cell state-dependent sQTLs identification, context-biased sQTLs discovery and colocalization with disease GWAS data.

### 2) Causal and null simulations to assess power and calibration

We evaluated the performance of *ISSAC* in null and causal settings to assess calibration and power, comparing *ISSAC* to LeafCutter^19^, currently the only method applied to map single-cell pseudobulk-level sQTL^15^. Because *ISSAC* and LeafCutter use different phenotype quantification, we simulated single-cell isoform expression with scIsoSim^43^ to ensure a fair comparison and minimize bias towards either phenotypic setup. First, we assessed both methods with null simulations, where no genetic variant is associated with splice site usage. To ensure robustness, we simulated lower (N=20) and higher (N=100) cell counts per individual. Both *ISSAC* and LeafCutter maintained well-calibrated p-values across various settings (**Fig. 2a**).

**Fig 2:**
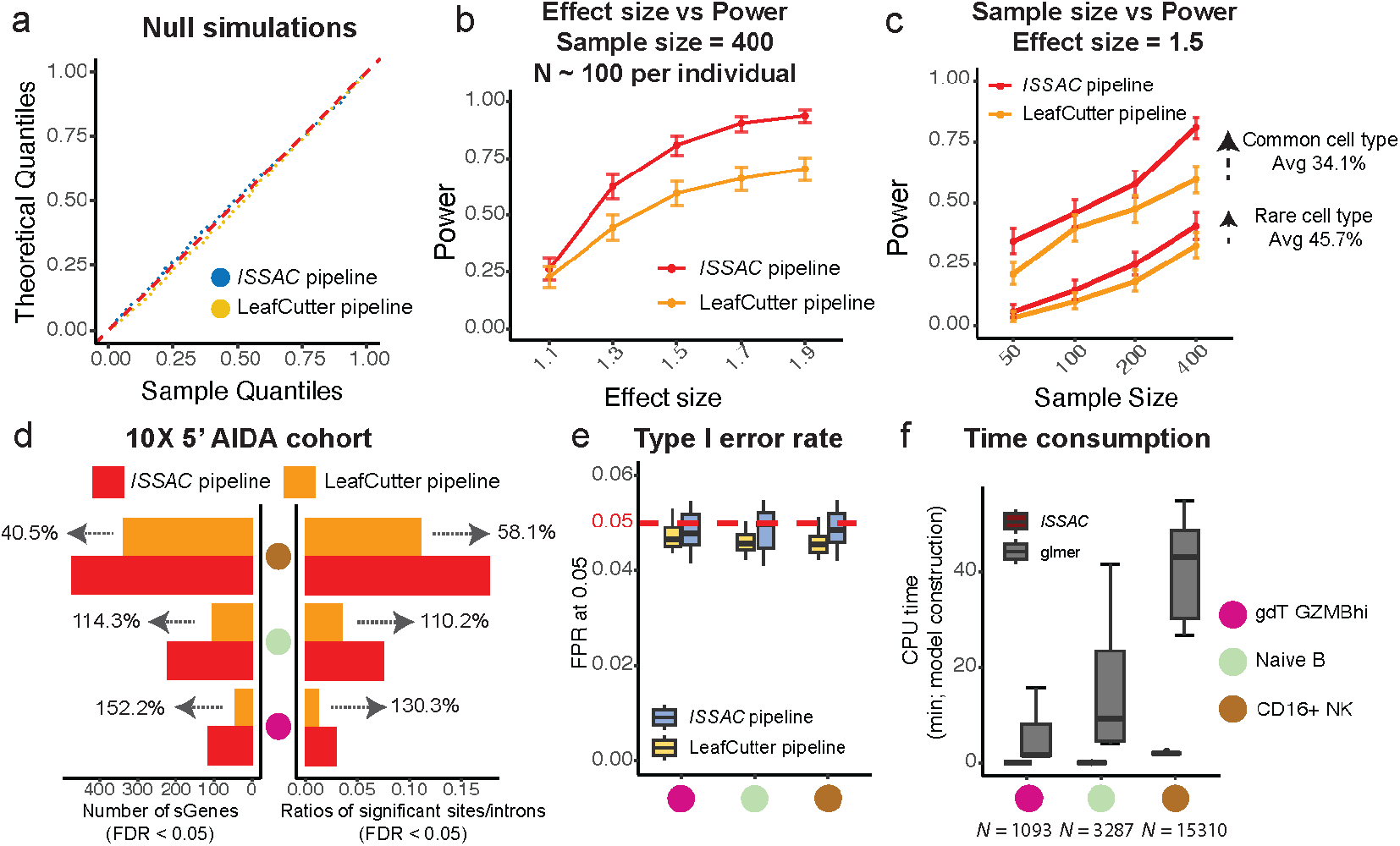
*ISSAC* exhibits higher power and proper calibration for sQTL mapping. **(a)** Calibration of *P* values for both *ISSAC* pipeline and LeafCutter pipeline when applied to splice site-based and intron-based phenotypes in the simulated scRNA-seq datasets; 1,000 random SNPs with MAF > 0.05 were generated to assess the calibration of both pipelines. **(b)** Power of *ISSAC* and LeafCutter pipeline across simulated scRNA-seq datasets with varying effect sizes (from 1.1 to 1.9). **(c)** Power of *ISSAC* and LeafCutter pipeline across simulated scRNA-seq datasets with different sample sizes (N = 50, 100, 200, 400; for common cell type, each individual has 100 cells and for rare cell type, each individual has 20 cells). **(d)** Performance comparison using 10X 5’ scRNA-seq data from three cell types (gdT GZMBhi, Naïve B and CD16+ NK) in the AIDA cohort (phase I freeze II). Analysis was restricted to genes where both splice sites and introns could be quantified, to ensure fair comparison between methods. **(e)** Type I error rates of *ISSAC* pipeline and LeafCutter pipeline in three cell types from AIDA where 50 splice sites (for *ISSAC*) and 50 splice introns (for LeafCutter) underwent 250 times of phenotype data permutations to create empirical null distributions by disrupting the potential associations between variants +/-1MB and splice phenotypes. **(f)** Runtime comparison between *ISSAC* and R package lme4 *glmer* function when applied to site-based phenotype across three cell types with varying sample sizes (gdT GZMBhi, Naïve B and CD16+ NK) from AIDA cohort.

Next, we evaluated *ISSAC* and LeafCutter in causal simulations where a subset of genes has genetically regulated isoform expressions. We used the same individuals and cells from the null simulation, and selected 334 genes as causal genes, whose isoform expressions are associated with genotypes. MAF of predefined causal SNPs was fixed at 0.5 to maximize statistical power, ensuring that observed differences in sensitivity reflect the underlying statistical models rather than allele frequency effects; full details are provided in **Supplementary Table 1**. We observed that *ISSAC* demonstrated 16.0-40.3% higher power than LeafCutter across a wide range of effect sizes (*β*=1.1 to 1.9) at a false discovery rate (FDR) < 0.05 (**Fig. 2b**). *ISSAC* also achieved 15.0-72.6% higher power than LeafCutter across a range of sample sizes (N=50 to 400; **Fig. 2c**). The power improvement is more pronounced in low-abundance cell types (N=20) than highly abundant cell types (N=100; **Fig. 2c**). Power increased consistently with sample size and sQTL mapping remained feasible even with 50 samples, where 5.7% and 34.4% of ground-truth sGenes were recoverable for rare and common cell types, respectively, at an effect size of 1.5. A breakdown of power by ground-truth alternative splicing event type, including power versus effect size and power versus sample size analyses, is provided in **Supplementary Fig. 2**. To confirm that this advantage is not specific to MAF = 0.5, we additionally evaluated *ISSAC* and LeafCutter across a range of MAF settings (MAF = 0.05, 0.1, 0.2, 0.3, and 0.4) and observed consistent power improvement for *ISSAC* in all conditions (**Supplementary Fig. 3**). To ensure robustness of our comparison, we selected a range of FDR threshold (0.01, 0.05, and 0.1) and observed consistent power improvement for *ISSAC* (**Extended Data Fig. 2a, b**). To understand the source of power improvement, we systematically tested the effect of phenotypic construction (splice site usage vs intron usage), distributional assumption (binomial vs Gaussian), and modeling assumption (pseudobulk with fixed effects vs metacell with mixed effects) through ablation studies, changing one variable while keeping the others constant. Splice site phenotypic construction improved the power by an average of 16.2% (two-sided paired t-test p-value=1.69×10^-3^, **Extended Data Fig. 2c**), binomial response variable improved the power by an average of 19.8% (two-sided paired t-test p-value=1.65×10^-3^, **Extended Data Fig. 2d**), and metacell with mixed effects improved the power by an average of 3.39% (two-sided paired t-test p-value=3.71×10^-2^, **Extended Data Fig. 2e**).

### 3) Assessing power and calibration using real-world datasets

To assess performance on a real-world dataset, we next applied *ISSAC* to the Asian Immune Diversity Atlas datasets (phase I freeze version II)^15,44^, for which single-cell sQTL was previously analyzed by LeafCutter at pseudobulk level^15^. We selected three cell types with low (gdT GZMBhi; N = 11,860; 39.14 cells / donor), medium (Naïve B; N = 36,778; 65.91 cells / donor) and high (CD16^+^ NK; N = 151,655; 259.68 cells / donor) cell counts, and generated 1,093, 3,287, and 15,310 metacells respectively. A uniform average of ∼10 cells per metacell was applied to ensure comparable metacell sizes across methods. To ensure fairer comparison between methods, we restricted analysis to the same set of splice junctions, then further limited to genes where both methods successfully generated splicing phenotypes, resulting in 2,103 tested genes for gdT GZMBhi, 1,957 for Naïve B, and 2,081 for CD16^+^ NK. The numbers of tested splice sites and genes are summarized in **Supplementary Fig. 4**. *ISSAC* identified 2.52-fold (116 vs. 46), 2.14-fold (225 vs. 105), and 1.41-fold (475 vs. 338) as many sGenes as LeafCutter (FDR < 0.05, two-pass filtering; **Fig. 2d** and **Supplementary Table 2**), with greater power gains observed in rarer cell types, consistent with our simulation results and further validated across ten cell types spanning a range of cellular abundances (Spearman’s *ρ* = −0.806, *P* = 0.0082; **Supplementary Fig. 5**). Breakdown of detected sGenes (non-intron-retention alternative splicing vs. intron retention) is shown in **Supplementary Fig. 6**. Through overlap analysis between *ISSAC* and LeafCutter, we found that 80.4%, 82.9%, and 88.5% of sGenes detected by LeafCutter were also identified by *ISSAC* in gdT GZMBhi, Naïve B, and CD16+ NK cells, respectively, demonstrating strong concordance (**Supplementary Fig. 7a**). Furthermore, splice sites detected by *ISSAC* showed greater statistical power and concordant effect sizes compared to paired introns detected by LeafCutter (**Supplementary Fig. 7b**,**c**).

To validate the analytical choices underlying these results, we benchmarked *ISSAC*’s metacell construction against SEACells^30^ across four combinations of metacell algorithm and normalization strategy; *ISSAC* with SCTransform normalization achieved significantly higher per-individual silhouette scores and sQTL mapping power than other combinations in most settings across all three cell types (Wilcoxon signed-rank test, *P* < 0.0001; **Supplementary Fig. 8**). SEACells performed poorly in the rare cell type setting, likely reflecting its design assumption of large metacell sizes. We further evaluated the sensitivity of sQTL power to the minimum metacell size parameter (m) by varying m across 2–20 (gdT GZMBhi, naïve B) and 10–100 (CD16+ NK). Power increased monotonically as metacell size decreased, plateauing near a metacell size-to-donor cell count ratio of approximately 1/5 to 1/4, and then declined substantially beyond this ratio (**Supplementary Fig. 9**), indicating that metacell size must balance pseudoreplication against signal dilution from sparse aggregation within individual metacells.

To assess calibration of *ISSAC* and LeafCutter, we selected 50 sGenes with varying expression sparsity (low, median, high) from each cell type and performing 250 times phenotype permutations to create empirical null distributions for sQTL. Both methods maintained well-controlled type I error rates below 5% at FDR < 0.05 (**Fig. 2e**). Quantile-quantile plots further confirmed well-calibrated *P*-value distributions across all sparsity levels for both methods (**Extended Data Fig. 3**). To further evaluate robustness, we extended the Type I error assessment to two additional significance thresholds (α = 0.01 and α = 0.005) and stratified results by MAF bin and splice site usage level. FPR remained well-controlled across all strata and thresholds, demonstrating that model calibration is robust to variation in both allele frequency and splice site prevalence (**Supplementary Fig. 10**).

To highlight the importance of incorporating a genetic relatedness matrix (GRM) into the binomial mixed model, we focused on splice sites with substantial inter-individual relatedness (τ > 0.5) across three cell types: gdT GZMBhi, Naïve B, and CD16+ NK (7, 14, and 72 sites, respectively). For each cell type, we simulated 10,000 null SNPs (MAF = 0.5) and tested their association with either the original splice site phenotype (uncorrected for sample relatedness, using two-sided t-test p-values) or relatedness-corrected residuals from *ISSAC* (using score tests). The uncorrected phenotype exhibited severe Type I error inflation across all three cell types, confirming that unmodelled sample relatedness substantially distorts association statistics (**Supplementary Fig. 11a**). In contrast, *ISSAC*’s mixed model effectively controlled this inflation, as further illustrated by representative splice sites in each cell type (**Supplementary Fig. 11b**).

To accommodate for ever-increasing cell counts and sample sizes, we implemented *ISSAC* in C++ and employed several techniques to improve runtime speed. For instance, we used preconditioned conjugate gradients (PCG) to avoid direct computation of the inverse of a large matrix^42^, as well as sparse GRM to reduce memory cost. *ISSAC* achieves 11 to 115-fold improvement in speed compared with the *glmer* function in R package *lme4* (**Fig. 2f**) and can map sQTLs for 1000 individuals with 1 million cells in 5 minutes per splice site using a single CPU thread (**Extended Data Fig. 4a**).

### 4) Mapping cell type-specific *cis*-sQTLs in DLPFC

The human brain exhibits exceptionally high transcriptomic diversity, characterized by extensive alternative splicing and diverse isoforms^45^. Although prior studies have explored the genetic regulation of splicing at the bulk tissue level^5,46-48^ and cell type-specific genetic regulation of gene expression^33^, cell type-specific genetic regulation of splicing in the brain remains unexplored. We applied *ISSAC* to identify cell type-specific genetic regulation of splicing in the dorsolateral prefrontal cortex (DLPFC) using the harmonized snRNA-seq resource from the FunGen-xQTL project^33,35,49^. After preprocessing and quality control (**Methods**), 3,177,748 cells of 722 specimens from 530 donors (192 donors shared across two resources, *CUIMC1* and *MIT*) with both snRNA-seq and whole-genome sequencing (WGS) data were retained for analysis (**Supplementary Table 3**). Following previous single-nucleus studies of the DLPFC, we grouped cells into seven major cell types, including excitatory neurons, inhibitory neurons, astrocytes, oligodendrocytes, oligodendrocyte progenitor cells, and endothelial cells (**Fig. 3a**). We further classified major cell types into 95 sub-cell types using an established pipeline^33^, retaining 67 sub-cell types with >3,000 cells for *cis*-sQTLs mapping (**Extended Data Fig. 6**). We aggregated single cells into metacells for the seven major cell types and 67 sub-cell types, generating 23,143 and 87,936 metacells, respectively (**Online Methods**). We used *ISSAC* to identify *cis*-sQTLs within +/-1-Mb window of the splice site for the seven major cell types and detected 20,071 and 2,823 *cis*-sQTLs for protein-coding and long noncoding RNA (lncRNA) genes respectively (FDR < 0.05), representing 29.3% of all tested genes (**Fig. 3b** and **Supplementary Table 4** and **5**). The remaining 8,424 sQTLs are associated with other biotypes, including pseudogenes, or could not be mapped to any annotated gene. In addition, we detected 6,576 and 2,778 *cis*-sQTLs for protein-coding and lncRNA genes (FDR < 0.05), respectively, for the 67 sub-cell types (**Fig. 3e** and **Supplementary Table 6** and **7**), with the remaining 7,507 sQTLs mapping to other biotypes or lacking gene annotation. Among these, 3,104 sSites (18.41%) were specific to subcell types despite lower sample size than major cell types, highlighting the cellular heterogeneity within the major cell types. One example of sub-cell specific sQTLs lies in *MAPK10*, which regulates amyloid-beta precursor protein signaling pathway during neuronal differentiation through phosphorylating APP and contributes to neuronal damage when dysregulated^50^. The lead SNP rs6844363 (C>T) is associated with the increased usage of the *MAPK10* exon1 donor splice site chr4:-:86359657 only significantly in Exc.10 (*P* = 3.23×10^-17^, *β* = 0.137) but not in excitatory neurons overall (*P* = 0.216, *β* = −0.017) or other subcell types such as Exc.9 (*P* = 0.196, *β* = −0.019; **Extended Data Fig. 8a**).

**Fig 3:**
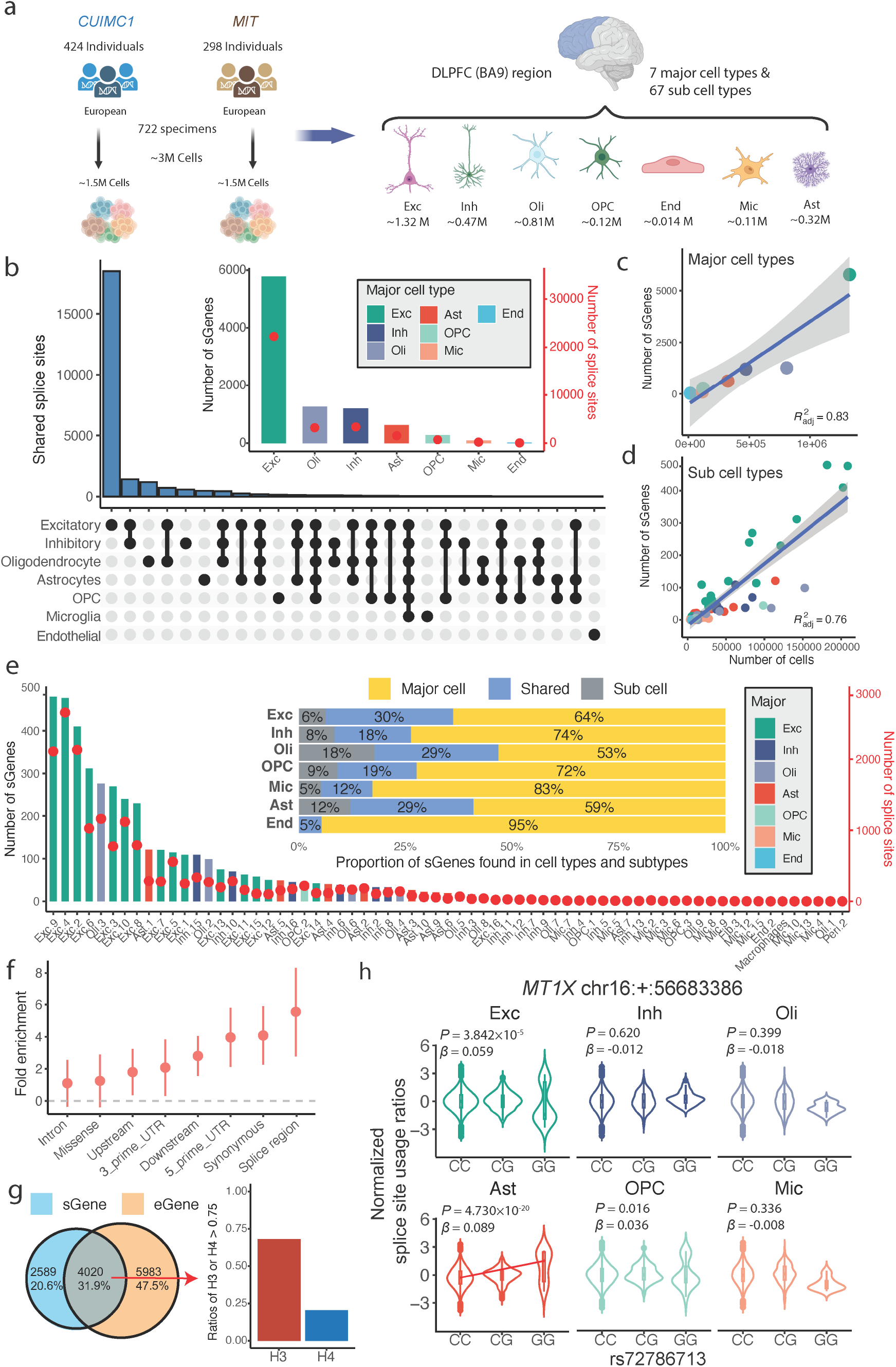
Major cell type level and subcell type level sQTL analysis revealed cell type-specific regulation of splicing. **(a)** Schematics of cell type-specific sQTL analysis in DLPFC(BA9) regions; 3 million cells from *CUIMC1* and *MIT* were integrated and divided into 7 major cell types & 67 subcell types for downstream sQTL analysis. **(b)** Numbers of sGenes and splice sites (FDR < 0.05) detected in the seven major cell types (top right); Numbers of shared significant splice sites between each major cell type (bottom left), each line connecting black dots represents one shared set. **(c)** Relationship between number of sGenes and number of cells in the 7 major cell types. The shaded area on either side of the linear regression line represents the 95% CI. **(d)** Relationship between number of sGenes and number of cells in the 67 subcell types. The shaded area on either side of the linear regression line represents the 95% CI. **(e)** Numbers of sGenes and splice sites (FDR < 0.05) detected in the 67 subcell types (bottom left); The proportion of sGenes shared by both major cell types and subcell types are indicated in the top right plot. **(f)** Fold enrichment of sSNPs in functional annotation category; the dashed line indicates the threshold of enrichment. **(g)** Sharing between sGenes and eGenes detected from seven major cell types; The distribution plot indicates the H_3_ and H_4_ of colocalized results between shared sGenes and eGenes. **(h)** Example of cell type-specific sQTLs; the lead SNP rs72786713 (C>G) is associated with the increased usage of the splice site chr16:+:56683386, with this effect being most prominent in astrocytes.

As a quality control, we observed that the number of significant sGenes positively correlated with the number of cells for both major cell types and sub-cell types (*R*^2^ = 0.83 and 0.76, respectively; **Fig. 3b-c**), as expected from statistical power considerations. We annotated the lead sSNPs for the 31,318 independent *cis*-sQTLs across the seven major cell types using SnpEff^51^ and estimated the fold enrichment of lead sSNPs in each functional category. The lead sSNPs were most enriched in the splice region (**Fig. 3f**), consistent with previous findings^15^. We performed replication analysis among the 192 shared individuals from the two resources, focusing on excitatory neurons. Storey’s π_1_ statistics showed high replication rate between them (π_1_ = 0.776 using *CUIMC1* as discovery and *MIT* as replication; π_1_ = 0.737 vice versa; **Extended Data Fig. 7**) as well as high correlation of effect sizes (Pearson’s *r* = 0.819 using *CUIMC1* as discovery and *MIT* as replication; Pearson’s *r* = 0.798 vice versa), suggesting that our *cis*-sQTLs were robust to batch effect and technical confounders.

To further compare *ISSAC* and LeafCutter, we performed LeafCutter-based sQTL mapping across seven major cell types, identifying 1,216, 302, 89, 72, 17, 7, and 1 sGenes in Excitatory neurons, Inhibitory neurons, Oligodendrocytes, Astrocytes, OPCs, Microglia, and Endothelial cells, respectively (**Supplementary Table 8**). To ensure comparability, analyses were restricted to genes testable by both methods. Among LeafCutter-detected sGenes, 93.0%, 89.3%, 92.7%, 81.6%, 66.7%, 100%, and 100% were also identified by *ISSAC* across the seven cell types. Beyond this overlap, *ISSAC* additionally detected 1,254, 245, 154, 86, 29, 13, and 2 sGenes (two-pass FDR < 0.05) not captured by LeafCutter, further demonstrating its improved sensitivity in 10X 3′ snRNA-seq data (**Supplementary Fig. 7d**). Restricting to the four major cell types with sufficient intron-site pairs for adequate comparison, *ISSAC* consistently yielded higher −log_10_(*P*)-values than LeafCutter (Wilcoxon signed-rank test: *P* = 1.77 × 10^-91^, 3.27 × 10^-15^, 8.74 × 10^-14^, and 1.59 × 10^-11^ in Excitatory neurons, Inhibitory neurons, Astrocytes, and Oligodendrocytes, respectively; **Supplementary Fig. 7e**), confirming greater statistical power in 10X 3′ snRNA-seq data. Effect size correlations between LeafCutter intron-based and *ISSAC* site-based sQTLs, using the splice-site lead SNP as reference, ranged from 0.679 to 0.789, indicating robust sQTL estimation by *ISSAC*.

Among all the 25,021 sSites (31,318 *cis*-sQTLs) that we detected across all cell types, 20,942 (83.7%) were detected to be significant in one of the seven major cell types, and 4,079 (16.3%) were detected to be significant in at least two cell types (**Fig. 3b**). Excitatory neurons yielded the most sQTL discovery due to their high abundances in the DLPFC. To uncover cell type-specific sQTLs, we used mash^52^ to quantify sQTLs in terms of sign and magnitude (**Supplementary Table 9**). We noted that sQTLs tend to share the same direction of effect but differ in magnitude across cell types (**Supplementary Fig. 12a**,**b**). One example of cell type-specific sQTLs is *MT1X*. The polymorphism of its lead sSNPs rs72786713 (C>G) led to increased usage of chr16:+:56683386 in the presence of the G allele, with the strongest effect observed in astrocytes (*P* = 4.730×10^-20^, *β* = 0.089, LFSR = 0, posterior mean = 0.089; **Fig. 3h, Supplementary Fig. 12c**). *MT1X* was reported to enable copper ion binding activity and the transcriptomic expression of *MT1X* increased along the temporal progression of AD in astrocytes^53^. This could help explain why *MT1X* splicing shows most prominent in astrocytes. An example of shared sQTLs is *PCM1*. The mutation of lead SNP rs2237848 (A>C) is associated with the increased usage of splice site chr8:+:17924712 in our six major cell types (**Extended Data Fig. 8b, Supplementary Fig. 12d**). *PCM1* was reported to be essential for centrosome assembly and function^54^, and plays a key role in cilia in maintaining brain structure and function^55^. It indicates that the splice regulation of *PCM1* plays a housekeeping role in almost all the neuron-related cells.

### 5) Distinct genetic regulation underlying cell type-specific sQTLs and eQTLs

We estimated the proportion of eQTL and sQTL effects that share the same underlying genetic regulatory mechanism. By comparing our sGenes to eGenes from an existing study using the same cohort^33^, we found that an average of 67.4% (50.5% to 88.0%) sGenes were not eGenes in their corresponding cell type. Among the overlapping sGene-eGenes, 20.6% have H_4_ > 0.75, indicating shared causal variants, while the majority of 68.8% have H_3_ > 0.75, indicating distinct regulatory variants (**Figure 3g**). We also compared sGenes detected by LeafCutter with eGenes from the same cohort. A substantial proportion of LeafCutter-detected sGenes were not captured by eGenes, ranging from 37.1% in Excitatory neurons to 100% in Endothelial cells (**Supplementary Fig. 13a**). Furthermore, the proportion of sGene-eGene pairs with H_3_ > 0.75 substantially exceeded those with H_4_ > 0.75 (**Supplementary Fig. 13b**), consistent with results obtained by *ISSAC*. Overall, our results indicate that the majority of cell type-specific sQTLs and eQTLs are regulated by distinct genetic mechanisms.

Among the sQTL-eQTL colocalization events, we further investigated potentially shared genetic regulatory mechanisms. Previous studies showed that alternative splicing, such as poison exon or intron retention, can lead to premature stop codon and nonsense-mediated decay^4,15,56,57^. A total of 298 colocalizing sQTLs observed in 230 sGenes regulated single-intron clusters, indicating potential intron retention events (**Fig. 4a**; **Supplementary Table 10**). In excitatory neurons, the acceptor splice site of *NAGK* exon 10 (chr2:+:71078317) was associated with the lead SNP rs6713 (C>T) (*P* = 4.24×10^-71^, *β* = −0.174; **Fig. 4b**), located 9 bp downstream of the splice site, and *NAGK* sQTLs-eQTLs colocalized with H_4_ = 0.99 (**Fig. 4c**). Retention of intron 9 introduces a Premature Termination Codon (TGA) at chr2:71,077,741–71,077,743, which lies 577 nucleotides upstream of the final exon–exon junction (chr2:71,078,318) in the *NAGK* mRNA — well exceeding the ∼50-nucleotide threshold^58^ required to trigger nonsense-mediated decay (NMD). This strongly supports the inference that intron 9 retention triggers NMD and likely leads to the observed downregulation of *NAGK* expression (**Fig. 4d**). *NAGK* is known for interacting with the dynein light chain roadblock type 1 to clear protein aggregates^59,60^. The downregulation of *NAGK* may impair its ability to clear protein aggregates and contribute to neurodegeneration. We extended this analysis to 72 intron retention sGenes with annotated splice sites that colocalized with eGenes and exhibited concordant sQTL/eQTL effect directions; all 72 (100%) harbored a PTC satisfying the 50-nucleotide rule (**Supplementary Table 11**), providing broad support for an NMD-mediated mechanism linking intron retention to reduced gene expression across these loci. We note that this analysis is restricted to annotated splice sites, as short-read sequencing data preclude full isoform reconstruction and thus prevent application of the 50-nucleotide rule at unannotated splice sites. Further, we replicated an intron retention event identified by isoMiGA project^61^. The lead SNP rs12006540 (T>C) modulates decreased usage of *HNRNPK* splice site chr9:-:83976994 in astrocytes (*P* = 1.7×10^-5^, *β* = −0.063; **Extended Data Fig. 8c**). The intron retention event near the site corresponds to the novel isoform characterized by the isoMiGA project using long-read RNA-seq. The independent corroboration by long-read sequencing provides orthogonal validation that *ISSAC*’s splice site usage ratio (SSUR)-based quantification can capture genuine intron retention events, including those involving unannotated isoforms that are inaccessible to intron-based methods.

**Fig 4:**
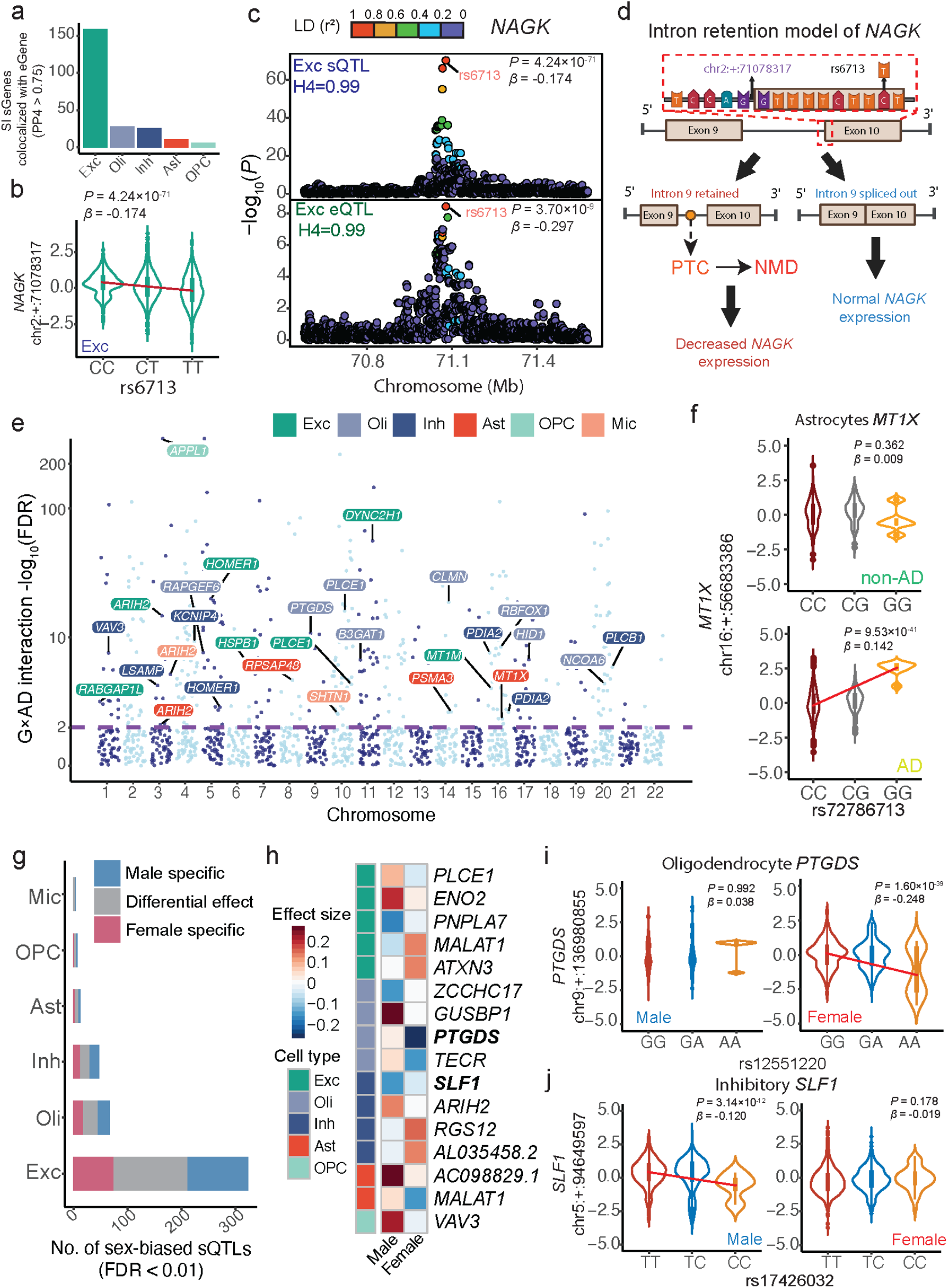
*ISSAC* revealed Intron retention sQTLs and context-biased sQTLs. **(a)** Numbers of sGenes discovered based on single intron clusters colocalizing with corresponding eGenes in major cell types (H_4_ > 0.75). **(b)** Violin plot showing the rs6713 (C>T) modulated the decrease usage of splice site chr2:+:71078317 within *NAGK*. **(c)** Colocalization event between *NAGK* eQTLs and sQTLs near the splice site chr2:+:71078317; rs6713 was the lead SNP for both eQTLs and sQTLs. **(d)** Intron retention model of *NAGK* showing rs6713 (C>T) potentially causes intron9 retention, leading to nonsense-mediated decay which further contributes to downregulation of *NAGK*. **(e)** Manhattan plots summarizing AD-biased sGenes in the major cell types. The FDR in the y axis −log_10_(FDR) represents the interaction term FDR between splice phenotype and the interaction term Genotype×AD. **(f)** Violin plot showing the associations between the lead SNP rs72786713 and splice site chr16:+:56683386 of *MT1X* in astrocytes is AD-biased. **(g)** Numbers of sex-biased splice sites in the major cell types (Genotype×Sex interaction FDR < 0.01). **(h)** Selected examples of sex-biased sGenes, with the color intensity reflecting sQTL effect sizes in males and females across five major cell types. **(i)** Violin plot showing the association between the lead SNP rs12551220 and splice site chr9:+:136980855 of *PTGDS* in oligodendrocyte is female-specific. **(j)** Violin plot showing the association between the lead SNP rs17426032 and splice site chr5:+:94649597 of *SLF1* in inhibitory neurons is male-specific.

### 6) Context-dependent splicing regulation in AD and sex

Splicing variants may influence disease susceptibility^62,63^, and conversely, disease status can alter the cellular and tissue microenvironment and modify genetic regulatory effects^36^. To identify sQTLs whose effects are modulated by Alzheimer’s disease (AD) pathology, we performed AD-biased sQTL analysis with a binomial mixed model implemented in the R package lme4^64^ to test for genotype-by-AD interaction. We identified a total of 399 AD-biased sQTLs in 194 sGenes across seven major cell types (Interaction FDR < 0.01; **Fig. 4e; Supplementary Table 12**). Notably, *MT1X*, previously identified as an astrocyte-specific sGene, shows strong AD-dependent regulation in astrocytes (**Fig. 4f**). The lead SNP rs72786713 (C>G) modulates the splice site (chr16:+:56683386) only in AD patients but not in healthy controls (control: *P* = 0.362, *β* = 0.009; AD: *P* = 9.53×10^-41^, *β* = 0.142). This aligns with previous reports of increased *MT1X* expression along the temporal progression of AD in astrocytes^53^, suggesting that AD pathology enhances sQTL effects on *MT1X*.

Similarly, to investigate sex-dimorphic splicing regulation, we performed sex-biased sQTLs detection with a binomial mixed model to test for genotype-by-sex (GxS) interaction. We identified 463 sex-biased sQTLs in 207 sGenes across seven major cell types (**Fig. 4g,h; Supplementary Table 13**). One example is that, in oligodendrocyte, the lead SNP rs12551220 (G>A) modulates the splice site chr9:+:136980855 within *PTGDS* specifically in females (**Fig. 4i**; male: *P* = 0.992, *β* = 0.038; female: *P* = 1.60 × 10^-39^, *β* = −0.248). *PTGDS* functions by converting prostaglandin H2 to prostaglandin D2 and plays a role in preventing the aggregation of amyloid-beta peptides, thereby conferring protection against Alzheimer’s disease^65,66^. Conversely, *SLF1* is an example of male-biased sQTLs. Within inhibitory neurons, the lead SNP rs17426032 (T>C) modulates the usage of splice site chr5:+:94649597 specifically in males (**Fig. 4j**; male: *P* = 3.14×10^-12^, *β* = −0.120; female: *P* = 0.178, *β* = −0.019). *SLF1* has been reported to participate in the DNA damage response pathway^67^, and has been implicated in two neurological disorders, form agnosia^68^ and Aicardi syndrome^69^.

Using available clinical covariates in the ROS/MAP cohort – specifically dcfdx (clinical diagnosis of cognitive status), cogdx (final consensus cognitive diagnosis), braaksc (neurofibrillary tangle burden), and ceradsc (neuritic plaque burden) – we tested the association between each interaction sQTL lead SNP and these clinical covariates using linear regression (clinical covariate ∼ genotype + age + sex). 17 AD interaction sQTLs and 16 sex interaction sQTLs associated with clinical covariates are prioritized (*P* < 0.01; **Supplementary Fig. 14a**,**b; Supplementary Table 14**,**15**). Specifically, the lead SNP rs8057953 (T>C), which reduces exon1 donor site usage at *MT1M* — a gene with neuroprotective roles^70^ (chr16:+:56632759; **Supplementary Fig. 14c**) — exhibits an AD-specific interaction: the splicing effect is significant in cognitively normal individuals (*β* = −0.108, *P* = 8.23 × 10^-18^) but absent in AD patients (*β* = −0.016, *P* = 0.449), and the same SNP associates with ceradsc (*β* = −0.580, *P* = 1.07 × 10^-4^), with the minor allele associated with reduced neuritic plaque severity. Similarly, the lead SNP rs190936155 (T>G), which increases exon3 donor site usage at *ZNF431* — encoding a KRAB-domain zinc finger transcription factor implicated in neuronal development^71^ (chr19:+:21151198; **Supplementary Fig. 14d**) — shows a female-specific splicing effect (female: *β* = 0.135, *P* = 1.78 × 10^-40^; male: *β* = 0.028, *P* = 0.239), and associated with both cogdx (*β* = 0.764, *P* = 9.69 × 10^-3^) and dcfdx (*β* = 0.943, *P* = 1.40 × 10^-3^), with the risk allele linked to worse cognitive diagnosis outcomes.

To compare metacell-level and donor-level pseudobulk approaches for interaction analyses, we performed genotype-by-AD and genotype-by-sex interaction analyses on significant sQTLs using both metacell-level and donor-level pseudobulk approaches. Metacell-level resolution yielded significantly higher statistical power than the donor-level approach in Excitatory neurons, Inhibitory neurons and oligodendrocytes for both genotype-by-AD (Wilcoxon signed-rank test, *P* = 1.4 × 10^-12^, *P* = 2.1 × 10^-3^ and *P* = 0.01 respectively, **Supplementary Fig. 15a, Supplementary Table 16**) and genotype-by-sex interactions (*P* = 7 × 10^-6^, *P* = 0.03 and *P* = 0.0041 respectively, **Supplementary Fig. 15c, Supplementary Table 17**). Among interaction sQTLs detected at FDR < 0.01 by the metacell-level approach, 56.6% (226/399) of AD-interaction and 58.1% (269/463) of sex-interaction sQTLs were replicated by the donor-level approach (**Supplementary Fig. 15b**,**d**). Together, donor-level pseudobulk data can detect a comparable number of interaction loci, likely benefiting from reduced within-donor metacell noise. At the same time, metacell-level resolution uniquely captures cell-state-dependent interaction effects whose signals are attenuated by donor-level aggregation and demonstrates significantly greater statistical power in neuronal populations.

### 7) Cell state-dependent sQTLs identification in inhibitory neurons

While cell type-specific sQTLs capture differences between distinct cell populations, regulatory effects can also vary across continuous cell states within the same cell type. Previous studies have shown that up to one-third of *cis*-eQTLs were influenced by multimodally defined cell states^23^. However, direct evidence of cell state-dependent genetic regulation of alternative splicing remains limited. Pseudobulk splicing quantifications, as we previously described^15^, could only detect mean cell-type effects and would mask cell state-dependent sQTLs. In contrast, metacells from *ISSAC* allowed us to capture continuous cell states using low-dimensional embedding from principal components analysis^72^ (PCA). Here, we focused on inhibitory neurons due to their relative abundance and elevated cellular diversity compared to other cell types^73^. Inhibitory neurons regulate brain activity by modulating the activity of excitatory neurons and play a critical role in synaptic plasticity^74^.

We scored each metacell along the top eight PCs and observed that each PC correlated with well-defined inhibitory neuron functions (**Fig. 5a**). Specifically, PC1 is enriched with neurogenesis genes based on gene set enrichment analysis (GSEA) against the Molecular signatures Database (MSigDB)^75,76^ (**Fig. 5b**). *ADARB2* and *SOX6* expression contributed most to PC1(Pearson’s *r* = 0.85 and −0.81, respectively). *ADARB2* is a key marker gene in central ganglionic eminence (CGE)-derived inhibitory neurons^77^, and its mutations have been linked to several neurological disorders^78-81^. *SOX6* is a gene that ensures proper formation and function of inhibitory neurons^82^. PC3 is enriched with synaptic signaling genes in GSEA results^76^ (**Fig. 5c**). *ERBB4* and *RALYL* expression were highly correlated with PC3 (Pearson’s correlation = −0.71 and 0.65, respectively). *ERBB4* has been reported to encode a receptor tyrosine kinase^83^ and promote inhibitory synapse formation^84^, and *RALYL* encodes an RNA-binding protein, and its decreased expression was correlated with the gradual progression of Alzheimer’s disease^85^.

**Fig 5:**
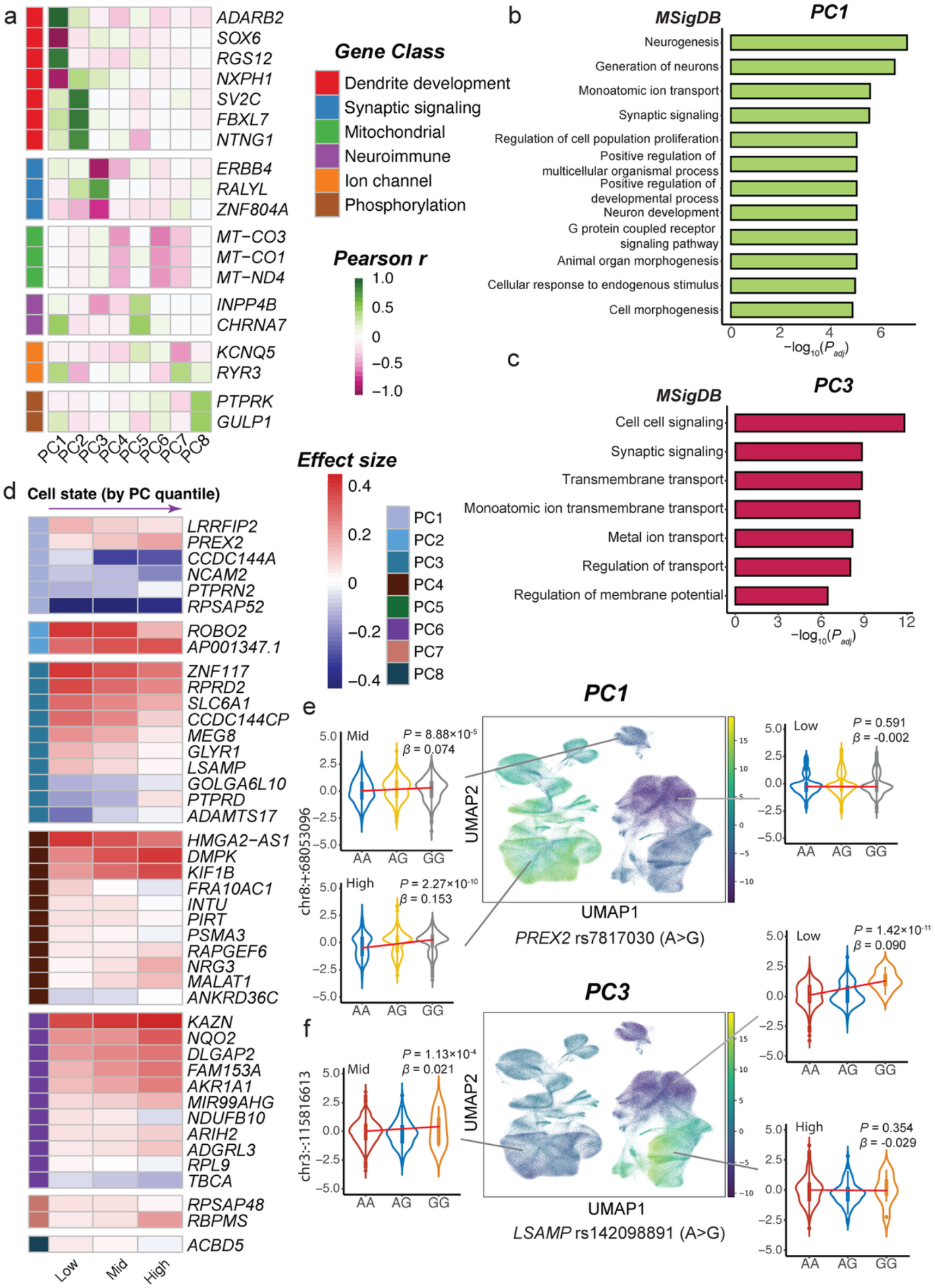
Cell state-dependent sQTLs identified in inhibitory neurons. **(a)** Heatmap showing Pearson’s correlations between PCs and selected marker genes’ normalized expression. **(b)** Summary of significant enrichment pathways identified by GSEA against MSigDB; Pathways are ordered by −log_10_(*P*_adj_); Genes were ranked by correlations between normalized gene expression and PC1 values. **(c)** Summary of significant enriched pathways identified by GSEA against MSigDB; Genes were ranked by correlations between normalized gene expression and PC3 values. **(d)** Examples of cell state-dependent sQTLs for PC-low (low 1/3 of PC values), PC-mid (mid 1/3 of PC values), PC-high groups (high 1/3 of PC values), respectively with the color bar representing cell states for PC1 to 8. **(e)** Uniform Manifold Approximation Projection (UMAP) plot of PC1 and one example of PC1-dependent sQTL *PREX2*; The color scale showing in the UMAP plot represents the PC1 values; Violin plots show the varied sQTL effect for cells in PC1-low (right), PC1-mid (left top) and PC1-high (left bottom) groups. **(f)** UMAP plot of PC3 and one example of PC3-dependent sQTL *LSAMP*; The color scale showing in the UMAP plot represents the PC3 values; Violin plots show the varied sQTL effect for cells in PC3-low (right top), PC3-mid (left) and PC3-high (right bottom) groups.

To map cell state-dependent sQTLs, we added genotype-by-cell-state (GxCS) interaction effect into the binomial mixed-effect model implemented in the R package lme4. We identified 369 *cis*-sQTLs to be cell state-dependent along the eight PCs in the major cell types (Interaction FDR < 0.05; **Fig. 5d; Supplementary Table 18**). Among them, *PREX2* sQTL showed interaction with PC1 (interaction *P* = 1.46 x 10^-7^). The genetic effect between lead SNP rs7817030 (A>G) and the splice site (chr8:+:68053096) in *PREX2* showed gradual increase along PC1 (**Fig. 5e**; PC1-low *P* = 0.591, *β* = −0.002; PC1-mid *P* = 8.88 x 10^-5^, *β* = 0.074; PC1-high *P* = 2.27 x 10^-10^, *β* = 0.153). *PREX2* has been reported to be involved in dendrite morphology and synaptic plasticity in Purkinje cells^86^, congruent with PC1’s representation of the dynamic changes of dendrite development. Another example is *LSAMP*; the effect size of its lead SNP rs142098891 (A>G) gradually decreased as PC3 increased (**Fig. 5f**; PC3-low *P* = 1.42 x 10^-11^, *β* = 0.090; PC3-mid *P* = 1.13 x 10^-4^, *β* = 0.021; PC3-high *P* = 0.354, *β* = −0.029). *LSAMP* is known to participate in the generation of a neuronal surface glycoprotein which acts as an adhesion molecule during axon guidance and synaptic formation^87^. Cell state-dependent splicing observed in *LSAMP* aligns with PC3’s role in the dynamic remodeling of synaptic organization. Other examples of cell state-dependent *cis*-sQTLs are shown in **Extended Fig. 9**. These results highlight the critical role that cell states play in the genetic regulation of splicing.

### 8) Linking cell type-specific sQTLs to complex neurological disorders

To understand cell type contributions to each disorder, we applied stratified LD score regression (S-LDSC)^88^ to estimate heritability enrichment of cell type-specific sGenes within GWAS summary statistics (**Fig. 6a**). We observed numerous cell type-specific enrichments across six GWAS (Enrichment Score > 1). For instance, microglia sGenes were highly enriched in AD and ALS GWAS but not for the other five traits. It has been observed that most AD risk genes were highly or exclusively expressed in microglia^89^, and activated microglia has been reported as a neuropathological hallmark of ALS^90^. Furthermore, oligodendrocyte sGenes were highly enriched in Parkinson’s disease. Oligodendrocyte-specific gene expression was reported to be distinctly associated with PD risk, but neuroinflammation-related cells, such as microglia, play a less causal role in PD^91^. As a quality control, we used BMI^92^ as negative controls and observed a lack of overall enrichment.

**Fig 6:**
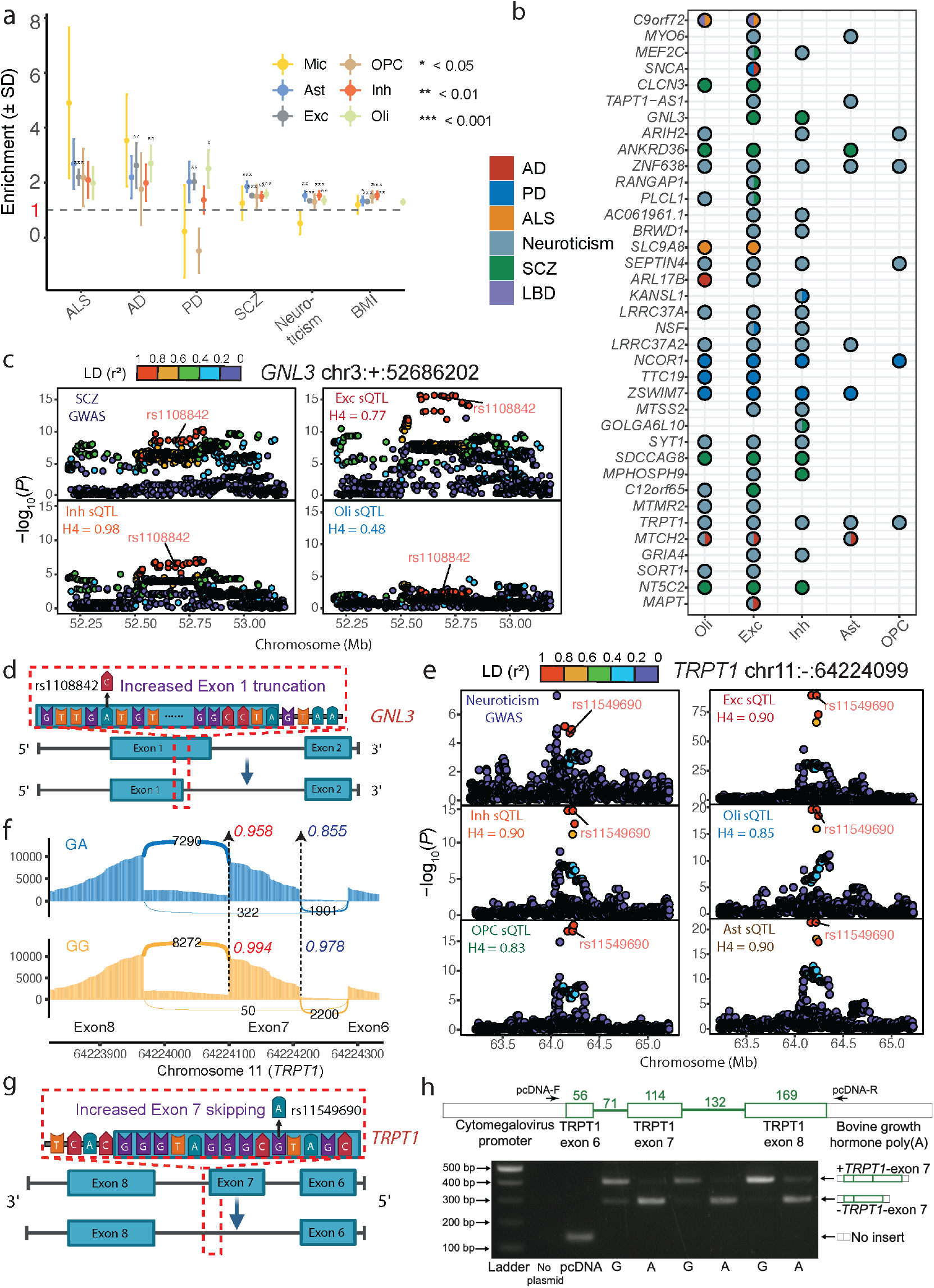
sQTLs detected by *ISSAC* revealed mechanism insights into neurological disorders. **(a)** Heritability enrichment (proportion *h*^*2*^/proportion variant) for five neuronal traits and one non-neuronal trait BMI mediated by *cis*-sQTLs from six major cell types (Exc, Inh, Oli, OPC, Ast and Mic). The dashed line represents the threshold for enrichment tendency within the trait GWAS for the significant sQTLs. **(b)** Colocalization events between significant sGenes with risk SNPs of AD, ALS, PD, SCZ, LBD and Neuroticism GWAS. The pie chart reports H_4_ over 0.75 of colocalization events detected by COLOC method. **(c)** Colocalization plots between SCZ GWAS and Exc, Inh, Oli sQTL around *GNL3* loci. **(d)** Illustration of possible mechanism how sSNP rs1108842 regulates the alternative donor splice site usage of exon1 within *GNL3*. **(e)** Colocalization plots between Neuroticism GWAS and Exc, Inh, Oli, OPC, Ast sQTL around *TRPT1* loci. **(f)** Sashimi plot demonstrates the exon skipping event modulated by the lead SNP rs11549690 (G>A) in *TRPT1*. The numbers within the introns represent UMI-based junction reads spanning the intron connecting exon6 and exon8; the red and purple numbers indicate exon7 donor and acceptor splice site usage ratio, respectively. **(g)** Illustration of possible mechanism how sSNP rs11549690 regulates the exon skipping events between exon6 and exon8. **(h)** Minigene experiment to validate the effect of rs11549690 on *TRPT1* exon 7 splicing in SH-SY5Y neuroblastoma cell line. The empty pcDNA3.1(+) backbone alone corresponded to the band with the smallest molecular weight on the gel image. The test region containing *TRPT1* exon 6, exon 7 and exon 8 was cloned into the pcDNA3.1(+) plasmid. Two identical minigene constructs with one nucleotide difference at rs11549690 (reference = G; alternative = A) were transfected into SH-SY5Y neuroblastoma cell line. The reference allele (G) predominantly led to the normal isoform with exon7 inclusion; the alternative allele (A) led to exon 7 exclusion.

To prioritize functional genes for complex neurological disorders, we performed colocalization analysis to assess whether cell type-specific sQTLs and trait are driven by the same underlying causal variants^93^. Here, we performed COLOC^93^ analysis between the seven major cell types and six well-powered neurological disorder GWAS including Alzheimer’s disease (AD), Amyotrophic lateral sclerosis (ALS), Parkinson’s disease (PD), Schizophrenia (SCZ), Lewy body dementia (LBD), and Neuroticism (**Supplementary Table 19**). We identified a total of 349 colocalization events across 142 sGenes with the six neurological diseases (H_4_ > 0.75; **Fig. 6b; Supplementary Table 20**). As an orthogonal validation, we cross-checked all colocalization events with summary-based Mendelian randomization (SMR)^94^, which replicated 310 of the 349 events (88.8%; *P*_*SMR*_ < 0.01; **Supplementary Table 20**). Of the 304 unique trait–sQTL locus pairs identified across five major cell types, a substantial proportion exhibit cell-type-specific colocalization (PP4 > 0.75 in the focal cell type and PP4 < 0.5 in all others), highlighting the functional heterogeneity of disease-relevant splicing across brain cell types (**Supplementary Fig. 16, Supplementary Table 21**). Notably, 41 loci colocalized exclusively in rarer cell types (oligodendrocytes, inhibitory neurons, astrocytes, or OPCs) but not in excitatory neurons, demonstrating that disease-associated splicing regulation is not solely driven by the most abundant cell type.

Among these findings, several colocalization events revealed particularly interesting insights. We identified a colocalization between AD and PD GWAS with an sQTL that regulates *MAPT* exon 3 inclusion in excitatory neurons (**Fig. 6b** and **Extended Data Fig. 10a, b**). *MAPT*, a well-known Alzheimer’s disease risk gene, encodes the microtubule-associated protein tau^95^. Here, the SNP rs17651213 (G>A) is a part of H1/H2 haplotype, increases the usage of splice site chr17:+:45974471 (*P* = 3.84 x 10^-13^; *β* = 0.081) and chr17:+:45974384 (*P* = 1.15 x 10^-9^; *β* = 0.061). It was previously reported that rs17651213 regulates exon 3 inclusion through common splice factors *hnRNPF* in a haplotype-specific manner^96^. Exon 3 encodes the N-terminal acidic projection domain that mediates the interaction of tau protein with cellular components implicated in AD and PD pathogenesis^97^. Further, we detected a colocalization event for *GNL3*, which produces a protein found within the nucleolus that binds p53 and plays an essential role in cell cycle and cell differentiation^98^. The colocalization between the *GNL3* sQTL and SCZ GWAS was most prominent in neuronal cell types, with strong evidence in inhibitory neurons (H_4_ = 0.98; *β* = 0.061) and excitatory neurons (H_4_ = 0.77; *β* = 0.084), but not in oligodendrocytes (H_4_ = 0.48; *β* = −0.004) (**Fig. 6c**). This cell-type-dependent pattern of colocalization probability may explain why this signal was not identified by previous bulk-level sQTL studies^5,8^. The lead SNP rs1108842 (A>C) mediated the splice site usage of chr3:+:52686105 (excitatory neurons: *P* = 2.09 x 10^-22^; *β* = 0.084; inhibitory neurons: *P* = 0.0001; *β* = 0.061; oligodendrocyte: *P* = 0.64; *β* = −0.004) and chr3:+:52686202 (excitatory neurons: *P* = 2.44 x 10^-16^, *β* = −0.077; inhibitory neurons: *P* = 2.29 x 10^-7^; *β* = −0.056; oligodendrocyte: *P* = 0.0431; *β* = −0.055) in a cell type-specific manner. The shift of *GNL3* exon1 donor splice site from chr3:+:52686202 to chr3:+:52686105 is associated with increased Schizophrenia risk (**Fig. 6d**). Other novel colocalization events, including *MTCH2* for AD and *TTC19* for PD, were reported in **Extended Data Fig. 10** and **Supplementary Table 20**.

We also identified a colocalization event between neuroticism and *TRPT1* sQTL, appearing in ectodermal cells including excitatory neurons, inhibitory neurons, OPC, oligodendrocyte, and astrocytes (**Fig. 6e**). *TRPT1* was predicted to enable tRNA 2’-phosphotransferase activity and catalyze the last step of tRNA splicing^99^. The lead SNP rs11549690 (G>A) modulated the usage of the exon7 donor splice site chr11:-:64224099 (excitatory neurons: *P* = 7.37 x 10^-89^, *β* = −0.180; inhibitory neurons: *P* = 3.89 x 10^-15^, *β* = −0.101; oligodendrocyte: *P* = 2.31 x 10^-20^, *β* = −0.139; OPC: *P* = 6.58 x 10^-18^, *β* = −0.167; astrocytes: *P* = 5.98 x 10^-22^, *β* = −0.177). The lead SNP rs11549690 is located 10 bp upstream of the splice site chr11:-:64224099. Switching from the reference (G) to alternative allele (A) increased exon-7 skipping read counts from 50 to 322 (6.4-fold; **Fig. 6f**).

To functionally validate our approaches, we focused on the colocalized *TRPT1* sQTL. To narrow down on the most likely causal variant, we used SpliceAI^100^ to predict the delta score induced by each variant. The lead variant was predicted to have the highest delta-donor score (Δ*Donor* = 0.21) on the splice site chr11:-:64224099, and we proceeded with functional validation to test the effect of rs11549690 on *TRPT1* exon 7 (114-nt) skipping in the SH-SY5Y neuroblastoma cell line. We designed a minigene construct whose test region consisted of the full sequences of *TRPT1* exons 6, 7 and 8 and intervening introns. We transfected two minigene constructs that are identical in sequence, except for a single nucleotide difference at rs11549690 (G or A), into SH-SY5Y cells. Both minigene constructs showed the presence of bands corresponding to exons 6 and 8 with either exon 7 inclusion (around 400-bp) or exon 7 exclusion (around 300-bp), but with the reference (G) allele showing a higher percentage of inclusion, and the alternative allele (A) showing a shift towards a higher percentage of exclusion (**Fig. 6g** and **h**). These functional validation experiments further support the causal effect of this SNP to the exon skipping events within *TRPT1*. These results highlight rs11549690 is a causal variant for *TRPT1* sQTL and potential contribution to Neuroticism risk via this exon skipping event. Overall, our findings provided potential target genes whose alternative splicing mediates complex neurological diseases at the single-cell level.

To assess whether colocalization events were missed by bulk and pseudobulk sQTL mapping, we compared *ISSAC*’s colocalization results against two reference datasets: GTEx v10 Brain Frontal Cortex (BA9) sQTL results and ROS/MAP pseudobulk sQTL results generated using the LeafCutter/QTLtools pipeline.

GTEx v10 Brain Frontal Cortex (BA9) sQTL results identified 52 colocalization events across AD, ALS, LBD, Neuroticism, and SCZ (**Supplementary Table 22, Supplementary Fig. 17a**). Against this reference, *ISSAC* identified 156 additional colocalized sGenes, of which 53.5% involved intron retention events undetectable by GTEx v10’s intron-based pipeline. For example, rs2779212 (T>C) was the lead SNP for an intron retention sQTL at *ZSWIM7* in excitatory neurons (chr17:−:15978043; *P* = 1.45 × 10^-39^, *β* = −0.12), colocalizing with a PD GWAS signal (*P* = 3.88 × 10^-8^, *β* = 0.089; H_4_ = 0.756; **Supplementary Fig. 17b**). *ZSWIM7* has previously been implicated as a PD risk gene^101^, and this finding provides a potential mechanistic link between aberrant alternative splicing at this locus and disease risk. Beyond intron retention, *ISSAC* also demonstrated finer splice-site resolution. Increased usage of the *G2E3* exon 2 acceptor (chr14:+:30581075) and donor (chr14:+:30581116) sites, driven by rs58049 (A>T; *P* = 1.50 × 10^-8^, *β* = 0.049 and *P* = 8.90 × 10^-12^, *β* = 0.075, respectively), colocalized with the ALS GWAS (H_4_ = 0.93) in excitatory neurons — an association absent from GTEx v10. Additionally, a subset of novel colocalizations reflects cell-type-specific sQTL effects undetectable in bulk tissue. Decreased usage of *MSANTD4* splice site chr11:−:106021835, driven by rs541640 (G>A), colocalized with the Neuroticism GWAS exclusively in astrocytes (H_4_ = 0.98) but not in excitatory neurons (H_4_ = 0.01, **Supplementary Fig. 17c**), underscoring the value of cell-type-resolved analyses for dissecting the cellular basis of neurological trait associations.

In the comparison with ROS/MAP pseudobulk sQTLs, the LeafCutter/QTLtools pseudobulk pipeline identified 83 colocalization events across 46 colocalized sGenes (**Supplementary Table 23**). In contrast, *ISSAC* uniquely identified 196 colocalized sGenes absent from pseudobulk results, representing 86.7% of all *ISSAC*-discovered colocalization events, while replicating 65.2% (30/46) of LeafCutter-detected sGenes — demonstrating strong concordance where both methods had sufficient power (**Supplementary Fig. 18a**). Beyond intron retention events inaccessible to LeafCutter (e.g., *ZSWIM7*/PD; **Supplementary Fig. 18b**), *ISSAC* showed greater sensitivity even for junction-based splice events in principle detectable by both methods. *ISSAC* detected sQTLs for increased usage of the *AUTS2* exon 2 donor site (chr7:+:69899498) and decreased usage of a novel splice site (chr7:+:69967804) in excitatory neurons, both driven by rs12698827 (chr7:69965762:C:T; *P* = 5.93 × 10^−7^, *β* = 0.057 and *P* = 6.97 × 10^−7^, *β* = −0.057, respectively), with strong Neuroticism GWAS colocalization (H_4_ = 0.90; **Supplementary Fig. 18c**). The corresponding LeafCutter intron-level sQTL (chr7:69899498:70118132:clu_13441_+) failed to colocalize with the same GWAS signal (H_4_ = 0.24), demonstrating that *ISSAC*’s splice-site resolution was essential for capturing this association. *AUTS2* is a well-established neurodevelopmental gene whose disruption underlies *AUTS2* syndrome^102^, encompassing autism spectrum disorder and intellectual disability.

## Discussion

We have developed *ISSAC*, a method to map cell type-specific and cell state-dependent sQTLs using sc/snRNA-seq data. Different from existing pseudobulk analysis^19^, *ISSAC* directly models splice site usages on the metacell-level using a binomial mixed-effect model. We showed via extensive simulation and real-world data that *ISSAC* was well-calibrated and detected 1.4-to 2.5-fold more sQTLs in diverse cell types compared to pseudobulk methods, with a greater power gain for low-abundance or rare cell types. Ablation studies demonstrated that *ISSAC* derived its power boost from a combination of splice site-based phenotypic construction, binomial distributional assumption, and metacell quantification with mixed-effect models. To accommodate for ever-increasing sample sizes, *ISSAC* was implemented in C++ with several modern techniques to improve runtime speed and achieved 11.2-to 115.1-fold faster speed than *glmer* function in the *lme4* R package.

We applied *ISSAC* to address the critical gap in identifying cell type-specific and cell state-dependent sQTLs in the human DLPFC, a brain region characterized by substantial isoform diversity and functional heterogeneity. More than 80% of sSites were unique to one of the seven major cell types, and thousands of sSites were identified only in subcell types and not in major cell types, highlighting the heterogeneity in splicing regulation. To address a key limitation of pseudobulk splicing quantification, we used *ISSAC* to uncover hundreds of cell state-dependent sQTLs in inhibitory neurons, revealing genetic effects that are modulated by dendritic development and trans-synaptic signaling. We prioritized 142 risk genes for neurological disorders, including *MTCH2, GNL3*, and *TRPT1*. We used minigene assays to functionally validate the genetic regulation mechanism of *TRPT1*. These results provide guidance for future investigations of drug targets for treating patients with neurological disorders at cell-level resolution.

Our results should be interpreted with their limitations, and we note several future directions of our work. First, the coverage of splice sites in our study is limited by the end-biased capture of mRNA molecules by droplet-based assays, which restricted our analysis to individual splice sites rather than full isoform reconstruction. Consequently, certain splicing events may not be detected by either *ISSAC* or LeafCutter pipeline since both methods were designed for local splicing quantification based on junctions. Isoforms that differ by the presence of additional exons at their 5’ or 3’ ends may be missed because splice sites at these terminal exons are typically connected to only one other splice site, preventing reliable quantification of their usage. Therefore, our analysis primarily captures internal alternative splicing events and may underestimate variation occurring at transcript termini. Emerging technologies, including combinatorial indexing with random priming^103^, scalable Smart-seq3xpress^104^, and single-cell long-read sequencing^105^, provides full-length transcript coverage. The adoption of these emerging methods is limited by protocol complexity, throughput, and cost, which are expected to improve over time. Another limitation of our framework is that splice site usage quantification generates multiple correlated phenotypes within a transcript. Because splice site usages are inherently compositional and statistically dependent, association testing at individual splice sites does not fully account for this correlation structure. Alternative strategies, such as modeling splice site usage using a multinomial response variable (e.g., as implemented in DRIM-seq^106^) or summarizing correlated splice site usage into a univariate representation in a reduced-dimensional simplex space (e.g., similar to sQTLSeeker^20^), may provide a more integrated statistical framework for association testing. Implementing and systematically evaluating these approaches may enhance the statistical modelling of splice site usage and represent promising directions for future methodological development. Third, our cell state analysis was restricted to inhibitory neurons and used principal components that, while capturing major axes of variation, may not correspond to specific biological states. Extending this analysis to other cell types and more interpretable cell state metrics (e.g., pseudotime, metabolic states) would provide deeper biological insights. Fourth, recent advancement in GPU computing has markedly reduced the runtime of QTL mapping software^107^. We believe the current CPU implementation of *ISSAC* could benefit from accelerated computing, and our GPU implementation of *ISSAC* is currently underway. Fifth, studies have shown that multi-ancestry QTL analysis would uncover additional ancestry-specific disease signals^15,16,23,44^. Our current cohorts primarily consist of donors with European ancestries, and additional donors with diverse genetic ancestry are critically needed in future recruitment. Lastly, our current list of prioritized disease genes will require extensive functional validation, and several validation efforts are underway.

## Methods

### Details of *ISSAC*

#### Step 1: splice read count and total read count quantification

*ISSAC* estimates splice site usage through UMI-collapsed junction reads as implemented in its *junctools* module. Different from bulk RNA-seq junction extraction tool which tracks read counts directly^108^, *ISSAC* collapses PCR-duplicate reads using both cell barcode and UMI to mitigate high PCR amplification biases in single-cell data. For a junction connecting upstream site *i* to downstream site *j*, we define collapsed read counts as *R*_*ij*_ = |*unique* (*barcode, UMI*) *pairs spanning junction* (*i, j*)|. Then for a target splice site *s*, we first identify its splice partners as the set of sites that form junctions with *s, P*(*s*) = {*p* : *R*_*sp*_ > 0 *or R*_*ps*_ > 0}. Junction reads supporting splice site *s* are those utilizing *s* as either upstream or downstream site, *Y*_*s*_ = ∑_*p*∈*P*(*s*)_(*R*_*sp*_ + *R*_*ps*_), while junction reads competing with *s* are those between partners of *s* that bypass *s* and do not form one isoform with one of the junctions utilizing *s*, junction reads crossing splice site *s* also taken into account, *C*_*s*_ = ∑_*p*∈ *P*_(_*s*_)_,)∉ *P*_(_*s*_)_,*q*≠*s*_(*R*_*pq*_ + *R*_*qp*_) + ∑_*p,q*∉ *P*_(_*s*_)_,*p*<*s,q*>*s*_ *R*_*pQ*_. The total reads count is therefore *N*_*s*_ = *Y*_*s*_ + *C*_*s*_. When *C*_*s*_ = 0, *i.e*., intron retention, unprocessed nascent transcript or incomplete junction detection due to 5’/3’ bias, we define *N*_*s*_ = *Y*_*s*_ + *R*′_*s*_ where *R*′_*s*_ denotes reads overlapping site *s* without splicing. It should be noted that intronic reads captured in 10x Chromium data may originate from either mature mRNAs retaining an intron or incompletely spliced pre-mRNAs, as the latter is known to be present in polyA-selected single-cell libraries. Therefore, our intron retention metric reflects intronic read coverage from both intron retention in mature mRNA and incomplete spliced pre-mRNAs and should be interpreted with this caveat in mind. These counts {*Y*_*s*_, *N*_*s*_} per metacell will be used in Step 2 to fit the binomial mixed model.

#### Step 2: fitting the null binomial mixed-effects model under the null hypothesis

Following splice site quantification, we fit a binomial mixed model under the null without genetic effects to estimate parameters for both fixed effects from covariates and variance components for random effects. For metacell *i* and splice site *s*, using the counts (*Y*_*is*_, *N*_*is*_) from Step 1, we construct a null association model without *cis*-sQTL effect as *logit*(*π*_*is*_) = *Xβ* + *b*, where *X* denotes fixed covariate such as sex, age and ancestry PCs for donors of metacell *i*, and *b* is the random effects accounting for metacell variability within donors. This model assumes *Y*_*is*_ ∼ *Binomial*(*N*_*is*_, *π*_*is*_) and *b* ∼ *N*(0, *τK*) where *K* is the genetic relationship matrix (GRM) capturing metacell and donor relatedness and *τ* represents the variance component.

Penalized quasi-likelihood (PQL) and REML methods are used to iteratively estimate the model parameters. Hereafter we drop the splice site subscript *s* to lighten notation. Inspired by SAIGE^31^ and fastGWA^109^, denote the quasi-likelihood of the observations conditional on the random effects as *ql*(*y* | *b*) and the log-likelihood of random effects distribution as *l*(*b*), the joint log quasi-likelihood is *ql*(*y* | *b*) + *l*(*b*) and marginal log quasi-likelihood, *ql*(*y*), can be computed as

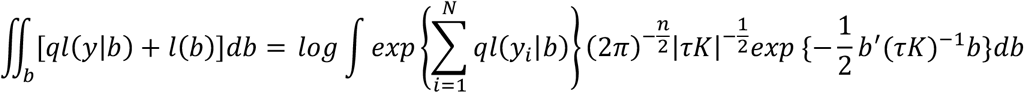

where 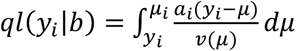 is the quasi-likelihood for the *i*th metacell when the random effect *b* is given and *a*_*i*_ is a constant which could be omitted.

To iteratively maximize *ql*(*y* | *b*) and *l*(*b*), we construct working variables at each iteration 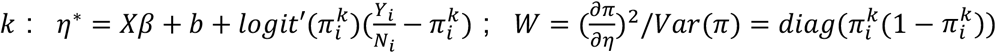; Here, the working response *η*^∗^ linearizes the binomial model around current estimates, and working weight *W* ensures observations with higher variance contribute less to parameter updates in order to account for heteroscedasticity. The variance of the working response is *Var*(*η*^∗^) = *W*^−1^ + *τK*.

Initializing with *τ* = 0, we estimate *β* and *b* by maximizing the REML pseudo-log-likelihood,

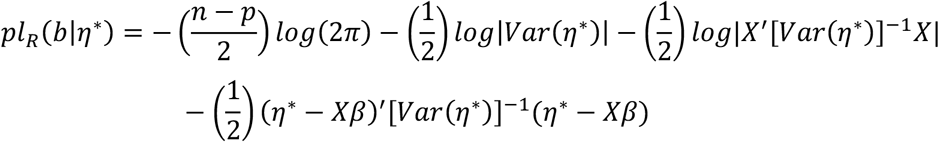

where *p* = *rank*(*X*) is a constant which could be ignored in the estimation process. The detailed estimation procedures are shown as following:

1. Obtain initial estimates of fixed effect *β* using iteratively reweighted least squares (IRLS) with *τ* = 0. The IRLS algorithm iterates over the following steps until convergence: (i) initialize *β* = 0 and the working response *η*^∗^ = *Xβ*; (ii) update predicted response 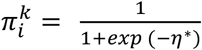; update working response 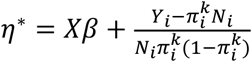; update working weights 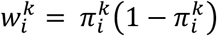; (iii) update fixed effect *β*_k+3_ = (*X*^T^W*X*)^−1^*X*^T^W*η*^∗^; (iv) Repeat (ii) and (iii) until convergence |*β*_*k*+3_ – *β*_*k*_| < *tol* (default tol: 0.001).
2. Update working variables using current estimates, for 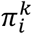, *η*^∗^, 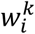.
3. Estimate variance components *τ* by maximize the REML pseudo-log-likelihood *pl*_R_(*b*|*η*^∗^) previously described, using the “opt.optimize” function from *nlopt* C++ library (https://nlopt.readthedocs.io/).
4. 4) Given estimated *τ*, update *β, b, Var*(*η*^∗^):

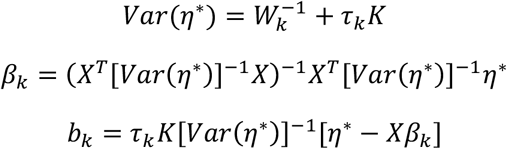

We repeat 2) to 4) until convergence |*τ*_*k*_ – *τ*_*k*−1_| < *tol* (Default: tol = 10^-6^)

#### Step 3: sQTL mapping

After obtaining *η*_*null*_ from above step, test statistics for sQTL mapping is computed as 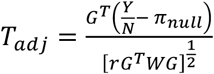 (**Supplementary Methods**), where 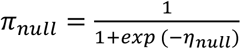 is predicted response under the null. *G* = *G*_*original*_ − *X*(*X*^*T*^*WX*)^−1^*X*^*T*^*G*_*original*_ is the covariate-adjusted genotype and normalization parameter *r* is obtained using a permutation method to adjust variance bias led by potential dispersion and random effects, by randomly sampling a certain number of genotypes with MAF > 0.05 (default: n = 1000). We obtained mean of observed variance through 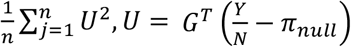. Then compute the expected variance through 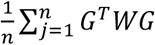. The parameter *r* is obtained through mean of observed variance divided by mean of expected variance.

Under the null, the test statistics is expected to follow a standard normal distribution after adjusting its variance bias through *r*. Corresponding *P* values can thus be obtained, and effect size be computed as the normalized regression slope between covariate-adjusted genotype and adjusted phenotype 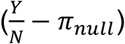.

##### Metacell calling procedure

To improve statistical power while maintaining low sparsity intrinsic to single-cell data, we generated phenotypes for downstream *cis*-sQTL based on the concept of metacells^110,111^. For each donor, we first normalized the raw count matrix using SCTransform^112^ which applies a regularized negative binomial regression to remove technical variation while preserving biological heterogeneity. We then clustered cells using PC embedding obtained from the SCTransform-normalized snRNA-seq gene expression matrix through *k* nearest neighbors’ graph construction^37^ and Louvain clustering^38^ (with *m* as the default parameters of the size of each cluster). Then we iteratively decomposed clusters with less than *m* cells to adjacent larger clusters to ensure each cluster having at least *m* cells. Finally, each cluster with at least *m* cells was treated as one metacell for downstream sQTL mapping tasks.

##### Simulation

To assess statistical performance of *ISSAC and* LeafCutter pipeline for sQTL discovery, we simulated 10x Genomics 5’ scRNA-seq data using scIsoSim^43^ and scReadSim^113^. To measure the performance under different effect size settings, we simulated five batches of sequencing reads with selected 334 protein coding genes (The five batches correspond to effect size 1.1, 1.3, 1.5, 1.7, 1.9). Each batch has 400 samples, with 100 cells per sample. For each gene, we simulated one ground-truth SNP and set the ratio of one isoform expression to scale linearly with SNP genotype at predefined effect sizes. Totally 334 predefined causal SNPs and 666 null SNPs were set for benchmark analysis (MAF = 0.5). To measure the performance under different sample size and number of cells settings for the two pipelines, two datasets with 400 samples were generated respectively (100 cells per individual for the common cell type dataset and 20 cells per individual for the rare cell type dataset; effect size = 1.5 for the two datasets). Then we subsampled the two datasets into 50, 100, 200 samples respectively to compare the performance under these sample size settings.

##### Benchmark studies

Here, we divided benchmark studies into three parts: Overall performance comparison, phenotype quantification benchmark and statistical model benchmark.

##### Overall performance comparison

We mapped 10x Genomics 5’ scRNA-seq fastq reads simulated previously using STARsolo^114^ and applied both *ISSAC* and LeafCutter pipeline including phenotype quantification and sQTL association testing. Power was defined as the proportion of ground-truth sGenes recovered at FDR < 0.05, where ground-truth sGenes were defined as genes harboring at least one significant splice site–predefined causal SNP pair. All splice sites and introns detected from the simulated scRNA-seq datasets were taken into consideration for the benchmark to ensure fairness.

##### ISSAC pipeline

We utilized metacell-level instead of pseudobulk-level as unit for sQTL mapping. Cells from the same individual were divided into two metacells averagely. Here, *ISSAC junctools* and *ISSAC juncstat* modules were firstly applied to obtain barcode- and UMI-based junction read counts. All the introns across 400 simulated samples were considered for splice site partner definition. *ISSAC pheno_group* module outputs *splice read count* and *total read count* which were used as phenotype for *cis*-sQTL mapping. As inter-individual genetic relationships were not modelled in the simulations, genetic relationship matrix was constructed with elements set to 1 if two corresponding metacells are from the same individual, and 0 otherwise. sQTL mapping was then performed using *ISSAC*’s binomial mixture model previously described for 1,000 simulated SNPs, with statistical significance set at FDR < 5% by applying the Benjamini-Hochberg (BH) procedure on the smallest nominal Pvalue of each splice site across all tested SNPs.

##### LeafCutter/QTLtools pipeline

*ISSAC junctools* and *juncstat* modules were used to obtain barcode- and UMI-based junction read counts for each pseudobulk-level donor. The *prepare_phenotype_table.py* script from LeafCutter was used to generate intron-based phenotype files in sQTL mapping. *cis*-sQTL mapping was performed using QTLtools v1.2, using intron excision ratios and 1,000 simulated SNPs. The threshold for statistical significance of splice introns was set at FDR < 5% by applying the BH procedure on the smallest nominal Pvalue of each intron across all tested SNPs.

##### Ablation study of phenotype quantification benchmark

To compare the performance of site-based quantification (*ISSAC*) and intron-based quantification (LeafCutter), we applied both methods to the simulated scRNA-seq datasets, following by a simple correlation analysis (cor.test function in R) to quantify associations between phenotype and predefined ground truth SNPs with the same multiple testing correction and power analysis procedures previously described.

##### Ablation study of statistical model benchmark

To measure the performance of different phenotype distribution assumption (Binomial vs. Gaussian) and different regression models (metacell with mixed effects model vs. pseudobulk with fixed effects model), the same site-based phenotypes were used for the three statistical models including *ISSAC* (Binomial mixed effects model), *ISSAC-fixed* (Binomial fixed effects model), QTLtools (Gaussian fixed effects model), under the same multiple testing correction and power analysis procedures previously described.

### ROS/MAP snRNA-seq data preprocessing

For this study, we collected 424 and 351 individuals’ 10x Genomics 3’ snRNA-seq datasets with corresponding WGS data focusing on DLPFC (BA9) regions from two studies^34,35^. Both datasets collected specimens coming from two longitudinal clinical pathological cohort studies which are the ROS and MAP^115^. Data for this study were obtained from AD Knowledge Portal under the request ID #12045. Gilad Sahar Green et al^34^ (referred to “*CUIMC1*” hereafter) and Hansruedi Mathys et al^35^ (referred to “*MIT*” hereafter) performed 10x Genomics 3’ snRNA-seq on donors coming from European descent. Data Generation has been detailed in previous study^34,35,49^. We annotated the cellular states of individual cells from each cell type separately based on the cell state definitions of *CUIMC1* to enable sQTL mapping combining two resources at the sub-cell level. The *CUIMC1* data and *MIT* data were first normalized, and the top variable genes were identified using the *SCTransform* function while regressing out percent of mitochondrial reads. A set of mapping anchors were identified and scored using the *FindTransferAnchors* function with default parameters. These anchors were then used to transfer the cell-state annotations from *CUIMC1* to *MIT* using the *TransferData* function. Removal of low-quality cells has been indicated in previous eQTL study^33^ and single-cell atlas of the aged human prefrontal cortex study^35^. After filtering specimens with few cells in major cell types (The threshold was set as following: endothelial (m = 5), OPC (m = 25), microglia (m = 25), astrocytes (m = 50), Inhibitory neurons (m = 50), oligodendrocyte (m = 100), Excitatory neurons (m = 100), m represents the minimum number of cells per donor), 424 and 298 specimens with corresponding genotype data were saved for downstream *cis*-sQTL mapping. The detailed information of all the 722 specimens is indicated in **Supplementary Table 3**. Finally, 3,177,748 cells were retained for downstream sQTL mapping tasks.

### Whole-Genome Sequencing

We leveraged publicly available data to identify 722 specimens with matched whole-genome sequencing (WGS) genotype data and single-nucleus RNA sequencing (snRNA-seq) data^116,117^. The WGS genotypes for 722 specimens were subset accordingly. Among the 722 specimens, 192 specimens coming from *CUIMC1* and 192 specimens coming from *MIT* belong to the same donors respectively. Thus the 530 unique individuals were involved in this study. We retained only valid biallelic single nucleotide polymorphisms (SNPs) and converted the genomic coordinates to the GRCh38/hg38 reference genome using CrossMap^118^. We estimated genetic sex based on X chromosome intensity ratios (F index) using PLINK(v1.9)^119^, defining males as F > 0.8 and females as F < 0.2. The genetic sex determination was used in downstream analyses as the sex variable. To ensure strand consistency, snpflip (https://github.com/biocore-ntnu/snpflip) was used, retaining only SNPs mapped to the forward strand. Phasing and genotype imputation were conducted using the Michigan Imputation Server^120^, with no additional QC performed on the pre-Imputation genotypes. The imputation utilized the 1000 Genomes Project Phase 3 high-coverage reference panel (GRCh38/hg38)^121^, incorporating data from all population groups. Following imputation, only variants with an imputation quality score of R^2^ > 0.8 were retained. Further filtering excluded imputed variants with MAF < 0.05, HWE *P*-values < 1×10^-6^, or non-biallelic variants. Quality control (QC) of the genotypes was performed using PLINK^119^. After completing all QC and filtering steps, a total of 5,781,824 SNPs were available for downstream QTL analysis.

### Demultiplex and Data preprocessing of ROS/MAP snRNA-seq datasets

In *CUIMC1*, each batch for library preparation consisted of eight participants except batch B63. Demultiplex (https://demultiplex.readthedocs.io/) was used to split the original fastq files consisting of multiple individuals into fastq files for each person. For *MIT*, the fastq files are already prepared for each person respectively. We used GENCODE release 32 (GRCh38, Ensembl 98, 5 September 2019) for gene annotation reference. snRNA-seq data were aligned to the human reference genome GRCh38 primary assembly and we utilized STARsolo v2.7.10a^114^ with the options – soloCBmatchWLtype 1MM –soloUMIdedup 1MM_Directional_UMItools to perform RNA mapping. The cell barcode whitelist was obtained from Cell Ranger installation (3M-february-2018.txt). A two-pass mode was used to enable new splice junction discovery and the waspOutputMode option was applied to reduce allelic mapping bias using VCF files from WGS data previously described. *ISSAC junctools* module enabled reads deduplication and UMI-based junction read counting.

### Mapping *cis*-sQTL in AIDA Data phase I Freeze v.2 dataset with *ISSAC and* LeafCutter/QTLtools pipeline

The AIDA Data phase I Freeze v.2 performed 10x 5’ scRNA-seq on 587 donors of Eastern (85 Singaporean Chinese, 148 Japanese, 165 Korean), Southeastern (60 Singaporean Malay, 59 Thai) and South Asian (70 Singaporean Indian) descent. The data generation and scRNA-seq preprocessing have been described in a previous study^15,44^.

Here, three cell types with varying cell counts were analyzed: gdT GZMBhi (11,860 cells), Naïve B (36,778 cells) and CD16+ NK (151,655 cells). To ensure comparison fairness, genes with both splice sites and intron data were taken into consideration, including 2,103 genes for gdT GZMBhi, 1,957 genes for Naïve B, and 2,081 genes for CD16+ NK.

#### *ISSAC* pipeline

We created metacells using the same procedure previously described for specimens having > 5 cells. Junctions extraction and phenotype construction procedures have been described previously (See in Methods part: Benchmark studies: *ISSAC* pipeline). *ISSAC pheno_output* module was used to filter sites with zero read counts in more than 50% of the samples or with insufficient variation (s.d. < 0.001). In terms of covariates, we considered 8 splice PCs, 5 genotype PCs, sex, age here. Genetic relationship matrix of the 587 donors was obtained through Plink *–make-grm-gz* option from genotype data^119^. Details of model construction have been mentioned in subsection “Benchmark studies” part. The associations between SNPs located within +/-1-Mb window of the splice site and splice site usage ratios were obtained through *ISSAC QTL* module. Here, we implemented two-pass multiple testing correction approach: in the first pass, Bonferroni correction is applied across all SNPs tested per splice site to identify the lead SNP (lowest Bonferroni-adjusted *P*-value) at each site; in the second pass, Benjamini–Hochberg FDR correction is applied across the list of lead SNP Bonferroni-adjusted *P*-value from all tested splice sites, with significance declared at FDR < 0.05.

#### LeafCutter/QTLtools pipeline

Details of junctions extraction and phenotype preparation have been described previously. We considered 8 splice PCs, 5 genotype PCs, sex and age as covariates. *cis*-sQTL mapping was performed using QTLtools v1.2 between SNPs located within +/-1-Mb window of the intron and intron excision ratios^122^, under the same two-pass multiple testing correction previously described.

### Mapping *cis*-sQTLs in ROS/MAP snRNA-seq dataset with *ISSAC*

We varied minimum cells per meta cell based on total cell counts of each cell type to balance between statistical power and data sparsity: 5 cells for endothelial, 25 for OPC, 25 for microglia, 50 for astrocytes, 50 for inhibitory neurons, 100 for oligodendrocyte and 100 for excitatory neurons. For subcell types, the minimum cell count thresholds were : N>200,000, m=50; 100,000<N<=200,000, m=30; 50,000<N<=100,000, m=15; 10,000<N<=50,000, m=10; N<=10,000, m=5 where N is cell numbers of the subcell type and m is the minimum cell numbers per meta cell. These thresholds were chosen such that the resulting metacell sample size is approximately 4-to 5-fold that of the corresponding pseudobulk sample size, a range empirically determined to maximize statistical power based on our metacell size optimization experiments (**Supplementary Fig. 8**). We then applied the *ISSAC* pipeline as previously described, taking into consideration introns with more than 20 UMI-based read counts across all samples. sQTL association testing were performed with several fixed effect covariates in addition to sex, age, genotypic and splice PC, including postmortem interval, clinical AD status, cohort (ROS or MAP), education years and number of cells per metacell. Two-pass multiple testing correction approach was implemented the same as in AIDA phase I freeze II cohort.

Leafcutter/QTLtools pipeline was additionally applied to the ROS/MAP snRNA-seq dataset for benchmarking pseudobulk-level sQTL mapping performance against *ISSAC*. Junction reads were extracted using regtools^108^, and intron clustering was performed with *leafcutter_cluster_regtools.py*. After quality control (minimum 50 junction reads per cluster; sparsity threshold < 50%), the following numbers of introns were retained per cell type: 39,729 (Excitatory neurons), 17,922 (Inhibitory neurons), 7,405 (Oligodendrocytes), 5,350 (Astrocytes), 2,315 (OPCs), 1,433 (Microglia), and 257 (Endothelial cells). The same covariate settings used in the metacell-level analysis were applied here. *cis*-sQTL mapping was performed using QTLtools v1.2 between SNPs located within +/-1-Mb window of the intron and intron excision ratios^122^, under the same two-pass multiple testing correction previously described.

Mashr v0.2.79^52^ was used to estimate sQTL sharing across cell types. Nominal P-values from sQTLs (sSites-sVariants) were used as input for each cell type. *Cis*-sQTLs were required to be significant (FDR < 0.05, two pass filtering) in at least one cell type and testable across six major cell types in the ROS/MAP cohort: Astrocytes, Excitatory neurons, Inhibitory neurons, Microglia, Oligodendrocytes, and OPCs. Endothelial cells were excluded due to an insufficient number of testable splice sites. A total of 2,676 sQTLs meeting these criteria were designated as the strong sets which were used to learn data-driven covariance matrices, and 10,000 randomly selected *cis*-sQTLs were used as the random sets which were used to learn correlation structures. The fitted model was used to compute the posterior means and LFSR for the strong sets.

### Enrichment of *cis*-sQTLs for functional annotations

We annotated lead *cis*-sQTLs detected by *ISSAC* in the 7 major cell types using SnpEff^51^ to test if *cis*-sQTLs were enriched in functional annotations. We sampled at random the same numbers of *cis*-SNPs from genotype data of this study as control SNPs. The sampling procedure was repeated 50 times to reduce stochastic biases. Then we computed the fold enrichment in each functional annotation category relative to mean frequencies of control SNPs across 50 replicates using in-house R scripts.

### AD- and sex-biased *cis*-sQTLs detection

AD- and sex-biased sQTLs were defined as those *cis*-sQTLs with significant genotype-by-AD or genotype-by-sex interaction effect. To avoid confounding, we balanced numbers of male and female samples when performing AD-biased sQTLs mapping and numbers of AD and non-AD samples when performing sex-biased sQTLs mapping. For each significant sQTL identified in previous sQTL analysis, a binomial mixed model was fitted using R lme4 package^64^ function *glmer* with family = “*binomial*” for the seven major cell types to test for genotype-by-AD or genotype-by-sex interaction while controlling the same set of covariates previously described. sQTLs with interaction FDR < 0.01 were defined as AD-biased or sex-biased sQTLs. Sex-specific sQTL was defined as significant in the discovery sex (*P* < 1×10^-3^) and nonsignificant in the other sex (*P* > 0.05). Differential effects were defined as those sQTL significant in both sexes but differed in their effect size. For donor-level AD-biased or sex-biased sQTL detection, a binomial fixed model was fitted for each cell type to test for genotype-by-context interaction while adjusting for known and unknown confounders.

### Cell state-dependent *cis*-sQTLs detection

Cell states for inhibitory neurons were determined via PCA using function *sc.pp.pca* from Python package scanpy^123^ on the 2000 highly variable genes in the snRNA-seq data of inhibitory neurons. Harmony^124^ was used to integrate inhibitory neurons from two data sources while removing batch effects. Here, cell state-dependent *cis*-sQTLs were defined as those *cis*-sQTLs with a significant genotype-by-PC interaction effect using PC1 to 8 representing eight distinct cell state axes. For each sQTL previously identified, we adopt the same procedure as previously described in the “**AD- and sex-biased *cis*-sQTLs detection**” part to test for genotype-by-PC interaction terms on all metacell samples while controlling known or unknown confounders. Additionally, we stratified 4,833 metacells in inhibitory neurons into PC-low (n=1,611), PC-mid (n=1,611) and PC-high (n=1,611) based on PC values and performed sQTL analysis within each group.

### Heritability enrichment in complex traits

We obtained publicly available GWAS summary statistics of 6 neurological disorders including AD, PD, ALS, SCZ, LBD and Neuroticism (**Supplementary Table 19**). BMI trait was used as negative control^92^. We estimated sGene enrichment in these traits using S-LDSC v1.0.1 with default parameters from the S-LDSC pipeline (https://github.com/bulik/ldsc/wiki/Partitioned-Heritability), where “sGene positive variants” is defined as those within a window of +-100kb around each sGene with at least one independent *cis*-sQTL.

### Colocalization analysis

Colocalization analysis was performed using COLOC^93^ between 7 major cell types sQTLs and 6 neurological GWAS using the coloc.abf function of the COLOC R package v5.2.3. H_4_ > 0.75 was set as the threshold for colocalization. The colocalization results were visualized using LocusCompare v1.0.0^125^. GTEx v10^8^ brain frontal cortex (BA9) *cis*-sQTL summary statistics were downloaded for benchmarking the colocalization results.

### Functional validation

To validate the effect of rs11549690 on *TRPT1* exon 7 splicing, we generated two versions of a minigene construct consisting of the full sequence of *TRPT1* exons 6, 7 and 8, with intervening introns differing at rs11549690 with G vs A allele. Both versions of the minigene were ordered as gene fragments, amplified by PCR, and cloned into an expression plasmid. These minigene constructs were then amplified by transformation into 10-β *Escherichia coli*, with overnight culture, followed by plasmid extraction and purification. The expanded minigene constructs were transfected into the SH-SY5Y neuroblastoma cell line and incubated for 72 hours. RNA was extracted, reverse-transcribed, amplified by RT-PCR, and analyzed by gel electrophoresis. Gene fragments were synthesized by GenScript, while all primers were synthesized by Integrated DNA Technologies. More detailed procedures are available in **Section 7 of the Supplementary Note**.

## Ethics & Inclusion statement

For ROS/MAP cohort, study was approved by an Institutional Review Board of Rush University Medical Center. For AIDA phase I freeze v2 cohort, all participants have been approved by local ethics review committees prior to study enrollment.

## Statistics and reproducibility

We performed sQTL analysis using *ISSAC* on a combined cohort of ROS/MAP snRNA-seq datasets consisting of 722 independent specimens belonging to 530 European individuals. In the binomial mixed model constructed by *ISSAC*, 8 splice PCs, 5 genotype PCs, sex, age, PMI, AD or not AD, ROS or MAP, education years, number of cells and cohort source were considered as covariates. All the statistical tests described in the article were two-sided. The filtering process resulted in the final sample sizes for each cell type as detailed in **Supplementary Table 3,4,6**. Standard quality control was applied to remove genetic variants in the association analyses (missingness > 0.05, MAF < 0.05 or Hardy-Weinberg equation *P* < 10^-6^). The experiments were not randomized, and the investigators were not blinded to allocation during the experiments. The scripts to reproduce the main results of the paper are available at github https://github.com/boxiangliulab/ISSAC.

## Data availability

The AIDA datasets are available via the HCA Portal (open access: https://data.humancellatlas.org/explore/projects/f0f89c14-7460-4bab-9d42-22228a91f185; managed access: https://data.humancellatlas.org/hca-bio-networks/genetic-diversity/datasets, https://explore.data.humancellatlas.org/projects/35d5b057-3daf-4ccd-8112-196194598893). ROS/MAP snRNA-seq raw sequence are available at Synapse (https://www.synapse.org/Synapse:syn31512863 & https://www.synapse.org/Synapse:syn52293442). Raw whole genome sequencing files are available at Synapse (https://www.synapse.org/Synapse:syn10901595). Access to the datasets is restricted and requires a data use certificate (DUC) to be submitted. Cell type summary statistics for significant sQTL are deposited in Zenodo (https://doi.org/10.5281/zenodo.17199088), with complete summary statistics available on Synapse as part of the FunGen-xQTL resource (https://www.synapse.org/Synapse:syn69670647). Note: referees will receive Synapse login credentials from the editor to access the Synapse repositories during peer review.

## Code availability

*ISSAC* software can be found at https://github.com/boxiangliulab/ISSAC, with *ISSAC* analysis pipeline and auxiliary codes used to reproduce the analysis in the manuscript available at https://github.com/boxiangliulab/ISSAC/tree/ISSAC_analysis. The pipeline depends on the following publicly available tools: SAMtools^126^ and BEDTools^127^.

## Acknowledgements

The results published here are in whole or in part based on data obtained from the AD knowledge portal. We thank the Asian Immune Diversity Atlas Network for data sharing. B.L. is supported by the Ministry of Education Singapore, under its Academic Research Fund Tier 2 (MOE-T2EP30123-0015), Academic Research Fund Tier 1 (FY2023; 23-0434-A0001; 22-5800-A0001), A*STAR under the Nucleic Acid Therapeutics Initiative (Award no.: H24J5a0066), and Usydney-NUS Ignition Grant (25-2107-A0001)

## Notes

### Competing Interest Statement

The authors have declared no competing interest.

### Summary of Updates

Here, I updated the author lists to make it more comprehensive.

## Reference

1. Scotti, M.M. & Swanson, M.S. RNA mis-splicing in disease. Nature Reviews Genetics 17, 19–32 (2016).

2. Cooper, T.A., Wan, L. & Dreyfuss, G. RNA and disease. Cell 136, 777–793 (2009).

3. Li, Y.I. et al. RNA splicing is a primary link between genetic variation and disease. Science 352, 600–604 (2016).

4. Liu, B. et al. Genetic analyses of human fetal retinal pigment epithelium gene expression suggest ocular disease mechanisms. Communications Biology (2019).

5. Qi, T. et al. Genetic control of RNA splicing and its distinct role in complex trait variation. Nat Genet 54, 1355–1363 (2022).

6. Wilkinson, M.E., Charenton, C. & Nagai, K. RNA Splicing by the Spliceosome. Annual Review of Biochemistry 89, 359–388 (2020).

7. Consortium, G. Genetic effects on gene expression across human tissues. Nature 550, 204–213 (2017).

8. Consortium, G.T. The GTEx Consortium atlas of genetic regulatory effects across human tissues. Science 369, 1318–1330 (2020).

9. Liu, B. et al. Genetic Regulatory Mechanisms of Smooth Muscle Cells Map to Coronary Artery Disease Risk Loci. The American Journal of Human Genetics 103, 377–388 (2018).

10. Garrido-Martín, D., Borsari, B., Calvo, M., Reverter, F. & Guigó, R. Identification and analysis of splicing quantitative trait loci across multiple tissues in the human genome. Nature Communications 12, 727 (2021).

11. Ota, M. et al. Dynamic landscape of immune cell-specific gene regulation in immune-mediated diseases. Cell 184, 3006–3021. e17 (2021).

12. Mu, Z. et al. The impact of cell type and context-dependent regulatory variants on human immune traits. Genome Biol 22, 122 (2021).

13. Martens, J.H. & Stunnenberg, H.G. BLUEPRINT: mapping human blood cell epigenomes. Haematologica 98, 1487–9 (2013).

14. Schmiedel, B.J. et al. Impact of Genetic Polymorphisms on Human Immune Cell Gene Expression. Cell 175, 1701–1715 e16 (2018).

15. Tian, C. et al. Single-cell RNA sequencing of peripheral blood links cell-type-specific regulation of splicing to autoimmune and inflammatory diseases. Nature Genetics 56, 2739–2752 (2024).

16. Perez, R.K. et al. Single-cell RNA-seq reveals cell type–specific molecular and genetic associations to lupus. Science 376, eabf1970 (2022).

17. Yazar, S. et al. Single-cell eQTL mapping identifies cell type–specific genetic control of autoimmune disease. Science 376, eabf3041 (2022).

18. Oelen, R. et al. Single-cell RNA-sequencing of peripheral blood mononuclear cells reveals widespread, context-specific gene expression regulation upon pathogenic exposure. Nature Communications 13, 1–15 (2022).

19. Li, Y.I. et al. Annotation-free quantification of RNA splicing using LeafCutter. Nat Genet 50, 151–158 (2018).

20. Monlong, J., Calvo, M., Ferreira, P.G. & Guigó, R. Identification of genetic variants associated with alternative splicing using sQTLseekeR. Nature Communications 5, 4698 (2014).

21. Zhao, K., Lu, Z.-x., Park, J.W., Zhou, Q. & Xing, Y. GLiMMPS: robust statistical model for regulatory variation of alternative splicing using RNA-seq data. Genome Biology 14, R74 (2013).

22. Rumker, L. et al. Identifying genetic variants that influence the abundance of cell states in single-cell data. Nature Genetics 56, 2068–2077 (2024).

23. Nathan, A. et al. Single-cell eQTL models reveal dynamic T cell state dependence of disease loci. Nature 606, 120–128 (2022).

24. Zhou, W. et al. Efficient and accurate mixed model association tool for single-cell eQTL analysis. medRxiv, 2024.05.15.24307317 (2024).

25. Macosko, E.Z. et al. Highly parallel genome-wide expression profiling of individual cells using nanoliter droplets. Cell 161, 1202–1214 (2015).

26. Cao, J. et al. Comprehensive single-cell transcriptional profiling of a multicellular organism. Science 357, 661–667 (2017).

27. Gierahn, T.M. et al. Seq-Well: portable, low-cost RNA sequencing of single cells at high throughput. Nature Methods 14, 395–398 (2017).

28. Dent, C.I. et al. A basic framework to explain splice-site choice in eukaryotes. Nature Communications 16, 8284 (2025).

29. Dent, C.I. et al. Quantifying splice-site usage: a simple yet powerful approach to analyze splicing. NAR Genomics and Bioinformatics 3, lqab041 (2021).

30. Persad, S. et al. SEACells infers transcriptional and epigenomic cellular states from single-cell genomics data. Nature Biotechnology 41, 1746–1757 (2023).

31. Zhou, W. et al. Efficiently controlling for case-control imbalance and sample relatedness in large-scale genetic association studies. Nat Genet 50, 1335–1341 (2018).

32. Aguzzoli Heberle, B. et al. Mapping medically relevant RNA isoform diversity in the aged human frontal cortex with deep long-read RNA-seq. Nature Biotechnology 43, 635–646 (2025).

33. Fujita, M. et al. Cell subtype-specific effects of genetic variation in the Alzheimer’s disease brain. Nature Genetics 56, 605–614 (2024).

34. Green, G.S. et al. Cellular communities reveal trajectories of brain ageing and Alzheimer’s disease. Nature 633, 634–645 (2024).

35. Mathys, H. et al. Single-cell atlas reveals correlates of high cognitive function, dementia, and resilience to Alzheimer’s disease pathology. Cell 186, 4365–4385.e27 (2023).

36. Biamonti, G. et al. Alternative splicing in Alzheimer’s disease. Aging Clinical and Experimental Research 33, 747–758 (2021).

37. Cover, T. & Hart, P. Nearest neighbor pattern classification. IEEE Transactions on Information Theory 13, 21–27 (1967).

38. Blondel, V.D., Guillaume, J.-L., Lambiotte, R. & Lefebvre, E. Fast unfolding of communities in large networks. Journal of Statistical Mechanics: Theory and Experiment 2008, P10008 (2008).

39. Zhou, H.J., Li, L., Li, Y., Li, W. & Li, J.J. PCA outperforms popular hidden variable inference methods for molecular QTL mapping. Genome Biology 23, 210 (2022).

40. Stegle, O., Parts, L., Durbin, R. & Winn, J. A Bayesian Framework to Account for Complex Non-Genetic Factors in Gene Expression Levels Greatly Increases Power in eQTL Studies. PLOS Computational Biology 6, e1000770 (2010).

41. Chen, H. et al. Control for Population Structure and Relatedness for Binary Traits in Genetic Association Studies via Logistic Mixed Models. Am J Hum Genet 98, 653–66 (2016).

42. Kaasschieter, E.F. Preconditioned conjugate gradients for solving singular systems. Journal of Computational and Applied Mathematics 24, 265–275 (1988).

43. Yan, G. University of California, Los Angeles (2025).

44. Kock, K.H. et al. Asian diversity in human immune cells. Cell 188, 2288–2306. e24 (2025).

45. Yeo, G., Holste, D., Kreiman, G. & Burge, C.B. Variation in alternative splicing across human tissues. Genome biology 5, 1–15 (2004).

46. Raj, T. et al. Integrative transcriptome analyses of the aging brain implicate altered splicing in Alzheimer’s disease susceptibility. Nature genetics 50, 1584–1592 (2018).

47. Takata, A., Matsumoto, N. & Kato, T. Genome-wide identification of splicing QTLs in the human brain and their enrichment among schizophrenia-associated loci. Nature Communications 8, 14519 (2017).

48. Li, Y.I., Wong, G., Humphrey, J. & Raj, T. Prioritizing Parkinson’s disease genes using population-scale transcriptomic data. Nature communications 10, 994 (2019).

49. Comandante-Lou, N. et al. PLXNB1 and other signaling drives a pathologic astrocyte state contributing to cognitive decline in Alzheimer’s Disease. bioRxiv, 2025.02.24.639868 (2025).

50. Kunde, S.-A. et al. Characterisation of de novo MAPK10/JNK3 truncation mutations associated with cognitive disorders in two unrelated patients. Human Genetics 132, 461–471 (2013).

51. Cingolani, P. et al. A program for annotating and predicting the effects of single nucleotide polymorphisms, SnpEff: SNPs in the genome of Drosophila melanogaster strain w1118; iso-2; iso-3. Fly (Austin) 6, 80–92 (2012).

52. Urbut, S.M., Wang, G., Carbonetto, P. & Stephens, M. Flexible statistical methods for estimating and testing effects in genomic studies with multiple conditions. Nature Genetics 51, 187–195 (2019).

53. Serrano-Pozo, A. et al. Astrocyte transcriptomic changes along the spatiotemporal progression of Alzheimer’s disease. Nature Neuroscience 27, 2384–2400 (2024).

54. Dammermann, A. & Merdes, A. Assembly of centrosomal proteins and microtubule organization depends on PCM-1. J Cell Biol 159, 255–66 (2002).

55. Monroe, T.O. et al. PCM1 is necessary for focal ciliary integrity and is a candidate for severe schizophrenia. Nature Communications 11, 5903 (2020).

56. Neil, C.R. et al. Poison exons: tuning RNA splicing for targeted gene regulation. Trends in Pharmacological Sciences 46, 264–278 (2025).

57. Ge, Y. & Porse, B.T. The functional consequences of intron retention: alternative splicing coupled to NMD as a regulator of gene expression. Bioessays 36, 236–243 (2014).

58. Nagy, E. & Maquat, L.E. A rule for termination-codon position within intron-containing genes: when nonsense affects RNA abundance. Trends in Biochemical Sciences 23, 198–199 (1998).

59. Ripon, M.K.H. et al. N-acetyl-D-glucosamine kinase binds dynein light chain roadblock 1 and promotes protein aggregate clearance. Cell Death & Disease 11, 619 (2020).

60. Islam, M.A., Sharif, S.R., Lee, H., Seog, D.-H. & Moon, I.S. N-acetyl-D-glucosamine kinase interacts with dynein light-chain roadblock type 1 at Golgi outposts in neuronal dendritic branch points. Experimental & Molecular Medicine 47, e177–e177 (2015).

61. Humphrey, J. et al. Long-read RNA sequencing atlas of human microglia isoforms elucidates disease-associated genetic regulation of splicing. Nature Genetics 57, 604–615 (2025).

62. Koch, L. Altered splicing in Alzheimer transcriptomes. Nature Reviews Genetics 19, 738–739 (2018).

63. Nikom, D. & Zheng, S. Alternative splicing in neurodegenerative disease and the promise of RNA therapies. Nature Reviews Neuroscience 24, 457–473 (2023).

64. Bates, D., Mächler, M., Bolker, B. & Walker, S. Fitting Linear Mixed-Effects Models Using lme4. Journal of Statistical Software 67, 1–48 (2015).

65. Zhou, Y. et al. Structure-function analysis of human l-prostaglandin D synthase bound with fatty acid molecules. Faseb j 24, 4668–77 (2010).

66. Kannaian, B. et al. Abundant neuroprotective chaperone Lipocalin-type prostaglandin D synthase (L-PGDS) disassembles the Amyloid-β fibrils. Scientific Reports 9, 12579 (2019).

67. Ryder, Emma L. et al. Structural mechanisms of SLF1 interactions with Histone H4 and RAD18 at the stalled replication fork. Nucleic Acids Research 52, 12405–12421 (2024).

68. Loui, P. A Dual-Stream Neuroanatomy of Singing. Music Percept 32, 232–241 (2015).

69. Ha, T.T. et al. Aicardi Syndrome Is a Genetically Heterogeneous Disorder. Genes (Basel) 14(2023).

70. de Vries, L.E. et al. Gene-expression profiling of individuals resilient to Alzheimer’s disease reveals higher expression of genes related to metallothionein and mitochondrial processes and no changes in the unfolded protein response. Acta Neuropathol Commun 12, 68 (2024).

71. He, Z., Cai, J., Lim, J.-W., Kroll, K. & Ma, L. A Novel KRAB Domain-containing Zinc Finger Transcription Factor ZNF431 Directly Represses Patched1 Transcription*. Journal of Biological Chemistry 286, 7279–7289 (2011).

72. Mackiewicz, A. & Ratajczak, W. Principal components analysis (PCA). Computers & Geosciences 19, 303–342 (1993).

73. Luo, C. et al. Single nucleus multi-omics identifies human cortical cell regulatory genome diversity. Cell genomics 2(2022).

74. Markram, H. et al. Interneurons of the neocortical inhibitory system. Nature Reviews Neuroscience 5, 793–807 (2004).

75. Ashburner, M. et al. Gene Ontology: tool for the unification of biology. Nature Genetics 25, 25–29 (2000).

76. Subramanian, A. et al. Gene set enrichment analysis: A knowledge-based approach for interpreting genome-wide expression profiles. Proceedings of the National Academy of Sciences 102, 15545–15550 (2005).

77. Martini, L., Bardini, R. & Di Carlo, S. Meta-Analysis of cortical inhibitory interneurons markers landscape and their performances in scRNA-seq studies. in 2021 IEEE International Conference on Bioinformatics and Biomedicine (BIBM) 253–258 (IEEE, 2021).

78. Gasparini, C.F. et al. Case-control study of ADARB1 and ADARB2 gene variants in migraine. The journal of headache and pain 16, 31 (2015).

79. Olofsson, I.A. et al. Genome-wide association study reveals a locus in ADARB2 for complete freedom from headache in Danish Blood Donors. Communications biology 7, 646 (2024).

80. Konki, M. et al. Peripheral blood DNA methylation differences in twin pairs discordant for Alzheimer’s disease. Clinical epigenetics 11, 130 (2019).

81. Lee, E. et al. Single-nucleotide polymorphisms are associated with cognitive decline at Alzheimer’s disease conversion within mild cognitive impairment patients. Alzheimer’s & Dementia: Diagnosis, Assessment & Disease Monitoring 8, 86–95 (2017).

82. Tagariello, A. et al. Balanced translocation in a patient with craniosynostosis disrupts the SOX6 gene and an evolutionarily conserved non-transcribed region. J Med Genet 43, 534–40 (2006).

83. Anton, E.S. et al. Receptor tyrosine kinase ErbB4 modulates neuroblast migration and placement in the adult forebrain. Nature Neuroscience 7, 1319–1328 (2004).

84. Luo, B. et al. ErbB4 promotes inhibitory synapse formation by cell adhesion, independent of its kinase activity. Translational Psychiatry 11, 361 (2021).

85. Zhang, Y., Wang, J., Liu, X. & Liu, H. Exploring the role of RALYL in Alzheimer’s disease reserve by network-based approaches. Alzheimer’s Research & Therapy 12, 165 (2020).

86. Donald, S. et al. P-Rex2 regulates Purkinje cell dendrite morphology and motor coordination. Proceedings of the National Academy of Sciences 105, 4483–4488 (2008).

87. Um, J.W. & Ko, J. Neural Glycosylphosphatidylinositol-Anchored Proteins in Synaptic Specification. Trends in Cell Biology 27, 931–945 (2017).

88. Finucane, H.K. et al. Partitioning heritability by functional annotation using genome-wide association summary statistics. Nat Genet 47, 1228–35 (2015).

89. Gao, C., Jiang, J., Tan, Y. & Chen, S. Microglia in neurodegenerative diseases: mechanism and potential therapeutic targets. Signal transduction and targeted therapy 8, 359 (2023).

90. Clarke, B.E. & Patani, R. The microglial component of amyotrophic lateral sclerosis. Brain 143, 3526–3539 (2020).

91. Agarwal, D. et al. A single-cell atlas of the human substantia nigra reveals cell-specific pathways associated with neurological disorders. Nature communications 11, 4183 (2020).

92. Yengo, L. et al. Meta-analysis of genome-wide association studies for height and body mass index in ∼700000 individuals of European ancestry. Hum Mol Genet 27, 3641–3649 (2018).

93. Giambartolomei, C. et al. Bayesian Test for Colocalisation between Pairs of Genetic Association Studies Using Summary Statistics. PLoS genetics 10, e1004383 (2014).

94. Zhu, Z. et al. Integration of summary data from GWAS and eQTL studies predicts complex trait gene targets. Nature Genetics 48, 481–487 (2016).

95. Strang, K.H., Golde, T.E. & Giasson, B.I. MAPT mutations, tauopathy, and mechanisms of neurodegeneration. Laboratory Investigation 99, 912–928 (2019).

96. Lai, M.C. et al. Haplotype-specific MAPT exon 3 expression regulated by common intronic polymorphisms associated with Parkinsonian disorders. Molecular Neurodegeneration 12, 79 (2017).

97. Dixit, R., Ross, J.L., Goldman, Y.E. & Holzbaur, E.L. Differential regulation of dynein and kinesin motor proteins by tau. Science 319, 1086–1089 (2008).

98. Romanova, L. et al. Critical role of nucleostemin in pre-rRNA processing. J Biol Chem 284, 4968–77 (2009).

99. Yang, X. et al. Structural and biochemical insights into the molecular mechanism of TRPT1 for nucleic acid ADP-ribosylation. Nucleic Acids Research 51, 7649–7665 (2023).

100. Jaganathan, K. et al. Predicting splicing from primary sequence with deep learning. Cell 176, 535–548. e24 (2019).

101. Yao, S. et al. A transcriptome-wide association study identifies susceptibility genes for Parkinson’s disease. NPJ Parkinsons Dis 7, 79 (2021).

102. Loberti, L. et al. AUTS2-related syndrome: Insights from a large European cohort. Genetics in Medicine 27, 101375 (2025).

103. Kuchina, A. et al. Microbial single-cell RNA sequencing by split-pool barcoding. Science 371, eaba5257 (2021).

104. Nasir, M.H. et al. Scalable blockchains—A systematic review. Future generation computer systems 126, 136–162 (2022).

105. Al’Khafaji, A.M. et al. High-throughput RNA isoform sequencing using programmed cDNA concatenation. Nature Biotechnology 42, 582–586 (2024).

106. Nowicka, M. & Robinson, M.D. DRIMSeq: a Dirichlet-multinomial framework for multivariate count outcomes in genomics. F1000Res 5, 1356 (2016).

107. Taylor-Weiner, A. et al. Scaling computational genomics to millions of individuals with GPUs. Genome biology 20, 228 (2019).

108. Cotto, K.C. et al. Integrated analysis of genomic and transcriptomic data for the discovery of splice-associated variants in cancer. Nature Communications 14, 1589 (2023).

109. Jiang, L. et al. A resource-efficient tool for mixed model association analysis of large-scale data. Nature Genetics 51, 1749–1755 (2019).

110. Baran, Y. et al. MetaCell: analysis of single-cell RNA-seq data using K-nn graph partitions. Genome biology 20, 206 (2019).

111. Persad, S. et al. SEACells infers transcriptional and epigenomic cellular states from single-cell genomics data. Nature biotechnology 41, 1746–1757 (2023).

112. Stuart, T. et al. Comprehensive Integration of Single-Cell Data. Cell 177, 1888–1902.e21 (2019).

113. Yan, G., Song, D. & Li, J.J. scReadSim: a single-cell RNA-seq and ATAC-seq read simulator. Nature Communications 14, 7482 (2023).

114. Kaminow, B., Yunusov, D. & Dobin, A. STARsolo: accurate, fast and versatile mapping/quantification of single-cell and single-nucleus RNA-seq data. (bioRxiv, 2021).

115. Bennett, D.A. et al. Religious Orders Study and Rush Memory and Aging Project. J Alzheimers Dis 64, S161–s189 (2018).

116. Hodes, R.J. & Buckholtz, N. Accelerating Medicines Partnership: Alzheimer’s Disease (AMP-AD) Knowledge Portal Aids Alzheimer’s Drug Discovery through Open Data Sharing. Expert Opinion on Therapeutic Targets 20, 389–391 (2016).

117. Leung, Y.Y. et al. Alzheimer’s Disease Sequencing Project release 4 whole genome sequencing dataset. Alzheimer’s & Dementia 21, e70237 (2025).

118. Zhao, H. et al. CrossMap: a versatile tool for coordinate conversion between genome assemblies. Bioinformatics 30, 1006–1007 (2014).

119. Purcell, S. et al. PLINK: a tool set for whole-genome association and population-based linkage analyses. Am J Hum Genet 81, 559–75 (2007).

120. Das, S. et al. Next-generation genotype imputation service and methods. Nature Genetics 48, 1284–1287 (2016).

121. Auton, A. et al. A global reference for human genetic variation. Nature 526, 68–74 (2015).

122. Delaneau, O. et al. A complete tool set for molecular QTL discovery and analysis. Nature Communications 8, 15452 (2017).

123. Wolf, F.A., Angerer, P. & Theis, F.J. SCANPY: large-scale single-cell gene expression data analysis. Genome Biology 19, 15 (2018).

124. Korsunsky, I. et al. Fast, sensitive and accurate integration of single-cell data with Harmony. Nature Methods 16, 1289–1296 (2019).

125. Liu, B., Gloudemans, M.J., Rao, A.S., Ingelsson, E. & Montgomery, S.B. Abundant associations with gene expression complicate GWAS follow-up. Nat Genet 51, 768–769 (2019).

126. Li, H. et al. The Sequence Alignment/Map format and SAMtools. Bioinformatics 25, 2078–2079 (2009).

127. Quinlan, A.R. & Hall, I.M. BEDTools: a flexible suite of utilities for comparing genomic features. Bioinformatics 26, 841–842 (2010).

